# Coupled Epidemiological and Wastewater Modeling at the Urban Scale: A Case Study for Munich

**DOI:** 10.1101/2025.09.25.25336633

**Authors:** Julia Bicker, Natalie Tomza, Karina Wallrafen-Sam, Nina Schmid, Andreas F. Hofmann, Sascha Korf, Alain Schengen, Jasmin Javanmardi, Andreas Wieser, Martin J. Kühn, Jan Hasenauer

## Abstract

Disease surveillance and epidemiological modeling are critical to guide public health interventions, but model performance depends on data availability and quality. While clinical reports often suffer from under-ascertainment and delays, wastewater-based surveillance (WBS) can rapidly capture community infection dynamics by detecting viral RNA from both symptomatic and asymptomatic cases. However, WBS data can be difficult to interpret. Here, we present a coupled model of infectious disease and wastewater dynamics designed for scalability to large cities. We calibrate the model to the first COVID-19 wave in the city of Munich and quantify how sampling protocols, precipitation infiltration, viral decay, normalization strategies, and intervention timing can shape the relationship between wastewater measurements and disease prevalence, thereby improving the interpretability and practical value of wastewater data for epidemiological decision-making. We find that when appropriate sampling, normalization, and analysis strategies are utilized, wastewater data can reveal changes in prevalence earlier than clinical reports and provide advance warning of upcoming increases in disease burden. Our results guide WBS design and integration into predictive early-warning systems. Our modeling framework is generalizable to other COVID-19-like pathogens to help enable robust, cost-efficient disease monitoring.

## Introduction

The COVID-19 pandemic as well as the growing risk of future infectious disease outbreaks [1] have underscored the need for both comprehensive disease surveillance and accurate predictive modeling to guide timely public health interventions. However, the reliability of model results is dependent on appropriate parameter selection, which is in turn constrained by the quality of the available data. Traditional epidemiological data sources have inherent limitations, with confirmed infections being affected by under-reporting and reporting delays [2, 3], while large longitudinal cohort studies are often resource-intensive to the point of infeasibility. In contrast, wastewater-based surveillance (WBS) has the potential to offer prompt and cost-efficient insights into population infection levels [4, 5] by detecting viral RNA shed by symptomatic and asymptomatic infected individuals alike. WBS applications include the monitoring of various pathogens [6] as well as drug use [7] and antimicrobial resistance [8]. Its capabilities were widely demonstrated during the COVID-19 pandemic through initiatives like Germany’s AMELAG project [9] and the Dutch equivalent [10], with various studies establishing that SARS-CoV-2 levels in wastewater closely correlate with clinical indicators such as reported case counts and test positivity rates [11, 12, 13, 14]. Utilizing wastewater data as a complement to traditional epidemiological indicators thus presents a promising avenue to improving the predictive accuracy of infectious disease models.

Despite its advantages, WBS is subject to significant sources of uncertainty, making it challenging to translate wastewater data into actionable epidemiological insights. For instance, questions persist about how best to design wastewater sampling protocols in order to account for the interplay between population density, human mobility, environmental variability, and sewer infrastructure [15, 16]. Wastewater measurements can also be affected by in-network viral degradation and sample dilution due to rain- and groundwater infiltration, while the effectiveness of different normalization strategies remains in need of further study [17]. By providing insights into how various confounding factors affect the relationship between underlying infection prevalence and wastewater measurements, mathematical modeling can help guide the interpretation of WBS data, so that it can be fully leveraged by predictive models and early warning systems [18, 19].

To provide reliably interpretable results, modeling approaches that utilize WBS data must incorporate the complex system of interactions among human mobility, meteorological factors, the natural history and transmission processes of the disease of interest, pathogen shedding and decay, sewer network characteristics, and the collection and analysis of wastewater samples (Fig. 1). However, most previously published modeling studies in this field consider only one factor in isolation. For example, studies by Hart and Halden [20] and Phan et al. [21] provide guidance on accounting for viral decay while one by Wang et al. [22] investigated optimal sampling location placement, but their modeling approaches did not account for rainfall or human mobility. Meanwhile, other dynamic modeling studies have leveraged wastewater data to estimate prevalence or incidence adjusted for under-reporting, or to forecast epidemic trajectories. For example, Pant et al. [23] and Mohring et al. [24] both used systems of ordinary differential equations to model SARS-CoV-2 prevalence alongside wastewater virus levels in their respective study regions, with similar goals of identifying the true scale of an outbreak and the timing of its peak. However, these studies’ use of deterministic models limits the insights they can provide into the mechanisms underlying trends in wastewater data, especially in highly stochastic scenarios such as low-prevalence settings. Nourbakhsh et al. [25] coupled an ordinary differential equation-based model of disease transmission with an advection-dispersion-decay model of sewer dynamics, but as the latter model was not spatial, this study still did not account for the infrastructural heterogeneity of real sewer networks. Finally, Ruiz et al. [26] combined disease and wastewater particle agent-based models (ABMs) to simulate the linked spread of an infectious disease and contaminated wastewater, with a particular focus on choosing appropriate temporal resolutions, but left complexities such as the effects of rainfall out of the scope of their exploratory study. These limitations were partially addressed by our previous work on an integrative model linking an ABM of infection dynamics to a hydrodynamic model of wastewater processes via a viral shedding model [27], but this study considered only a small synthetic neighborhood over a short two-week simulation period, with applications to real-world scenarios left for future work.

**Figure 1:**
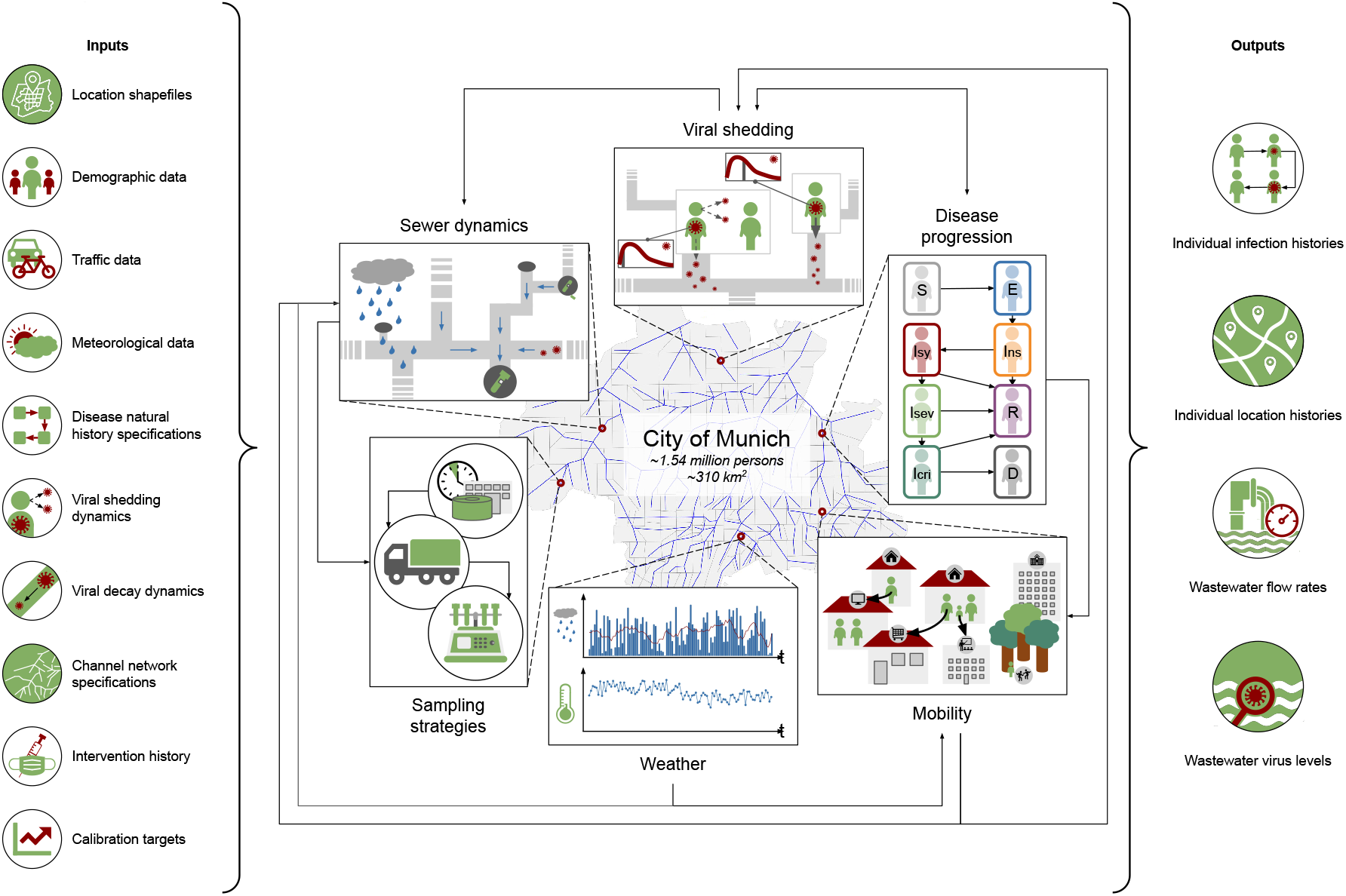
Overview of factors shaping disease spread and wastewater signals. Infection and wastewater dynamics are influenced by interconnected epidemiological, environmental, and infrastructural processes, informed by data from diverse sources (left panel). Human *mobility* shapes *sewer dynamics* via domestic wastewater inflow and modulates the effects of *viral shedding* by controlling where and near whom individuals shed. Fecal *viral shedding* into wastewater affects *sewer dynamics*, while respiratory *viral shedding* drives new infections, which each follow a *disease progression* pathway. *Disease progression*, in turn, alters *mobility* by prompting hospitalizations or NPIs. *Weather* affects *sewer dynamics* via rainwater infiltration and can also influence *mobility*. Finally, *sewer dynamics* determine the outcomes of wastewater surveillance *sampling strategies* (central panel). Together, these processes produce the observable outcomes of infection histories, location histories, wastewater flow rates, and wastewater virus levels (right panel).

In this study, we present an urban-scale modeling framework that integrates extensive heterogeneous data sources on demographics, mobility patterns, weather, infrastructure, topography, seroprevalence, reported cases, and public health intervention history to capture coupled infection and wastewater dynamics with high fidelity. We extend our previously developed mechanistic models of infectious disease, viral shedding, and sewer dynamics to enable novel simulations of an entire city over a three-month time horizon. As a case study, we calibrate the model to closely reflect the city of Munich, Germany during the first wave of the COVID-19 pandemic, from the beginning of March to the beginning of June 2020. Through a set of realistic counterfactual scenarios simulated using this framework, we investigate where and why disease “hotspots” emerge and explore the effects of sampling location, rainwater infiltration, viral decay, and human fecal indicator-versus flow-based normalization strategies on the relationship between true infection prevalence and wastewater measurements. We further demonstrate how non-pharmaceutical interventions (NPIs) such as mask-wearing or social distancing can temporarily alter this relationship. We thereby provide practical recommendations to public health practitioners on interpreting wastewater monitoring results, refining WBS systems, and advancing the development of predictive infectious disease models informed by wastewater data.

## Results

### Infection dynamics model explains heterogeneous transmission through realistic mobility

To assess the scalability of the infection dynamics ABM when integrating novel real-world data, we initially conducted a simulation study on this model in isolation. The ABM initialization step defined agents that realistically represent the population of Munich, with assigned locations reflecting real-world demographic and infrastructural patterns (Fig. 2a). Locations were mapped to agents based on trip data from a transport demand model called DEMO [28], which contains about 4.8 million trips, generated based on a mobility survey of Germany from 2017 [29, 30], for 1.3 million modeled persons living in Munich. Each trip includes individual-level information such as age and household identifier, as well as spatially explicit origins, destinations, and activity types, which were mapped to the ABM location types *Home, School, Work, Recreation*, and *Shop*. Except for locations outside of Munich assigned to commuting residents, all locations lie within DEMO traffic cells that divide the area of Munich into smaller disjoint areas [31], enabling subsequent mapping to wastewater catchment areas. Because the official population of Munich was roughly 1.5 million in 2022 [32], the closest census year to the modeled time frame, we scaled up the DEMO-derived population accordingly by sampling age, household size, and assigned locations from the underlying distributions (see Methods for details). While new households were created, each representing a new *Home* location, agents were assigned to existing locations of each non-*Home* type. The maximum number of contacts at *Work* and *School* locations were set to 40 and 45, respectively, based on reports from the POLYMOD social contact study [33]. We used German hospital registry data [34, 35] to generate *Hospital* and *ICU* locations.

**Figure 2:**
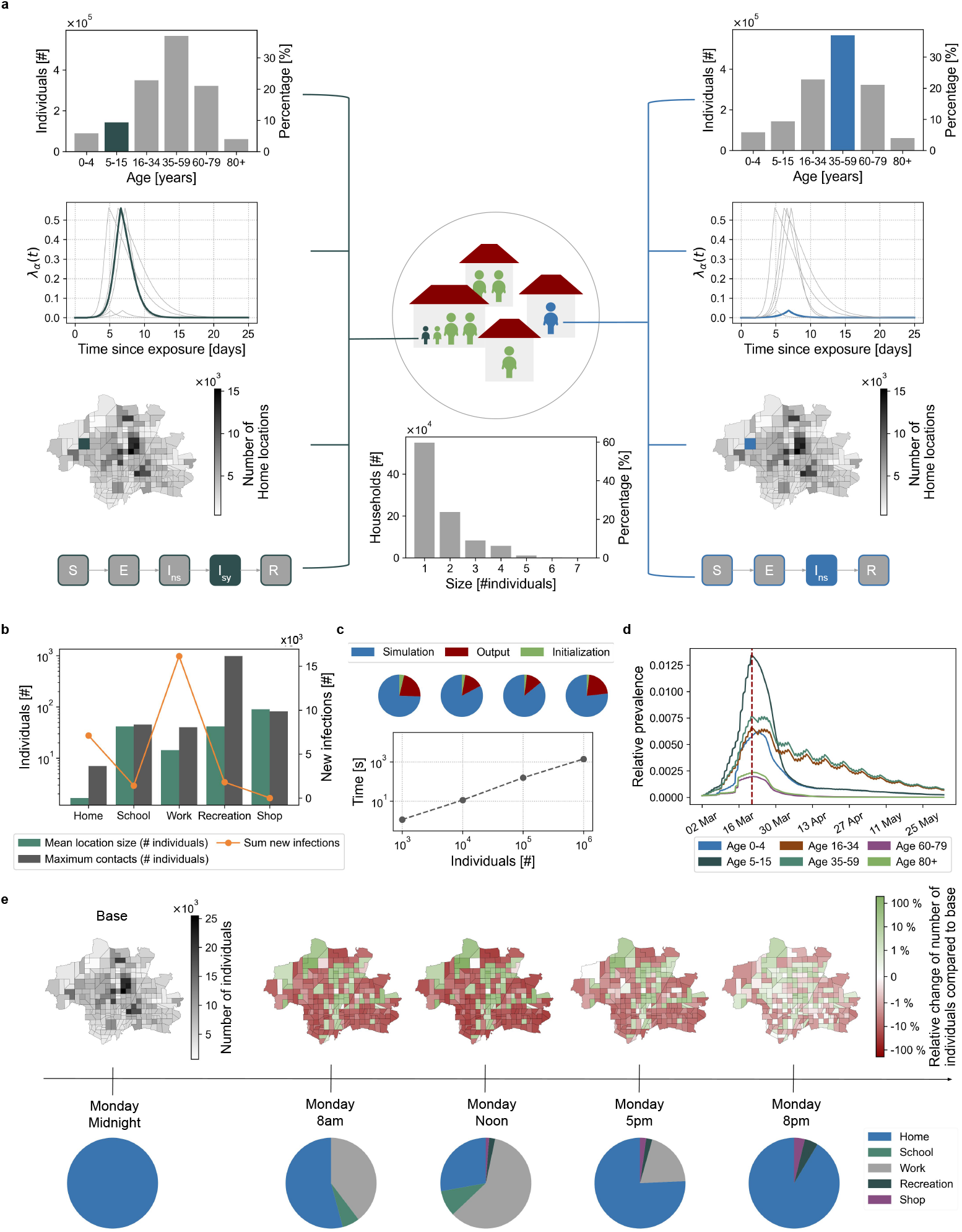
Overview of the infection dynamics model. **a)** Overview of the household structure in the ABM and a selection of attributes for two agents, shown in dark green and blue. The agent-dependent disease course can include the infection states Susceptible (*S*), Exposed (*E*), Non-symptomatically Infected (*I*_*ns*_), Symptomatically Infected (*I*_*sy*_), and Recovered (*R*), among others. **b)** Mean location sizes, maximum number of contacts, and total number of new infections during the simulated time frame for different location types. Shown are averages across 100 simulations. The majority of infections occur at *Work* (60%) or at *Home* (27%). **c)** ABM runtime scaling: Computational complexity scales linearly with the simulated number of agents. Hardware details are provided in Supplementary Table S2. **d)** Relative prevalence over time for every age group. Shown are the mean values across 100 simulations. The red line indicates start of lockdown on the nineteenth day. **e)** Movement of agents between wastewater areas and location types throughout the day, for one example simulation with a totally susceptible population.

The final initialized model contains about 1.5 million agents and about 1.0 million locations (88.4% *Home, <*1% *School*, 6.1% *Work*, 3.5% *Recreation*, 1.6% *Shop*, 30 *Hospital*, and 17 *ICU*) of varying size (Fig. 2b). Of these, roughly 95% fall within the wastewater catchment areas (see Supplementary Table S1 for exact numbers). Consequently, during periods spent outside the city, agents neither contribute wastewater nor shed fecal material into the city sewer system, and therefore have no influence on the wastewater model.

Based on official COVID-19 reporting standards [36], agents were stratified into six age groups: 0-4 years, 5-15 years, 16-34 years, 35-59 years, 60-79 years and 80+ years, leading to household-size-dependent age group distributions (Supplementary Fig. S1a-c). The second age group consists of schoolage agents, while the third and fourth comprise working-age agents, with daily trips to *School* and *Work* locations during working hours strongly influencing the population’s overall mobility behavior (Supplementary Fig. S1d).

To ensure computational feasibility, interfaces connecting the ABM to the wastewater and the shedding model were streamlined (Supplementary Fig. S2), and initialization and input/output routines were implemented in C++ for efficiency. The ABM runtime scales linearly with both population size and simulation time frame, with the simulation itself accounting for the majority of the computational cost, while initialization overhead is negligible (Fig. 2c).

The ABM simulates agent movement patterns across locations of different types, with observed variations driven by agent age group as well as by NPIs. For this simulation study, NPIs were implemented in the form of location closures and transmission rate dampings, which reduce the number of contacts at specific location types and the transmission rate of infectious individuals, respectively. For this study, the simulation time frame extended from 02 March 2020 to 01 June 2020, inclusive. Starting on Saturday, 21 March 2020 (when the first full lockdown began in Bavaria, Germany [37]) all *School, Shop*, and *Recreation* locations and 30% of *Work* locations [38] in the ABM were closed down; they then remained closed for the rest of the model time frame. A set of 100 simulations, with parameters estimated as discussed in the calibration results section, indicate that contact behavior and transmission dynamics differ substantially between location types (Fig. 2b, Supplementary Fig. S3a), which in turn influences prevalence heterogeneity across age groups (Fig. 2d) and household sizes (Supplementary Fig. S3b). Although school-age children are at an elevated risk of infection due to the large number of contacts they have at *School* (Fig. 2b,d), the largest number of infections occurs at *Work* (Fig. 2b) as the majority of the population is working-age. The model also produces dynamic changes in population density across wastewater catchment areas throughout the day (Fig. 2e, Supplementary Fig. S1e), thereby capturing spatial variability in water usage and fecal shedding.

### Wastewater models can be reconstructed from publicly accessible data

To assess the relation of the spread of an infection to the measured wastewater levels, we reconstructed the real-world sewage system of Munich using a geographic information system (GIS) [39] integrating publicly accessible hydrological, infrastructural, and topographic data (Fig. 3a).

**Figure 3:**
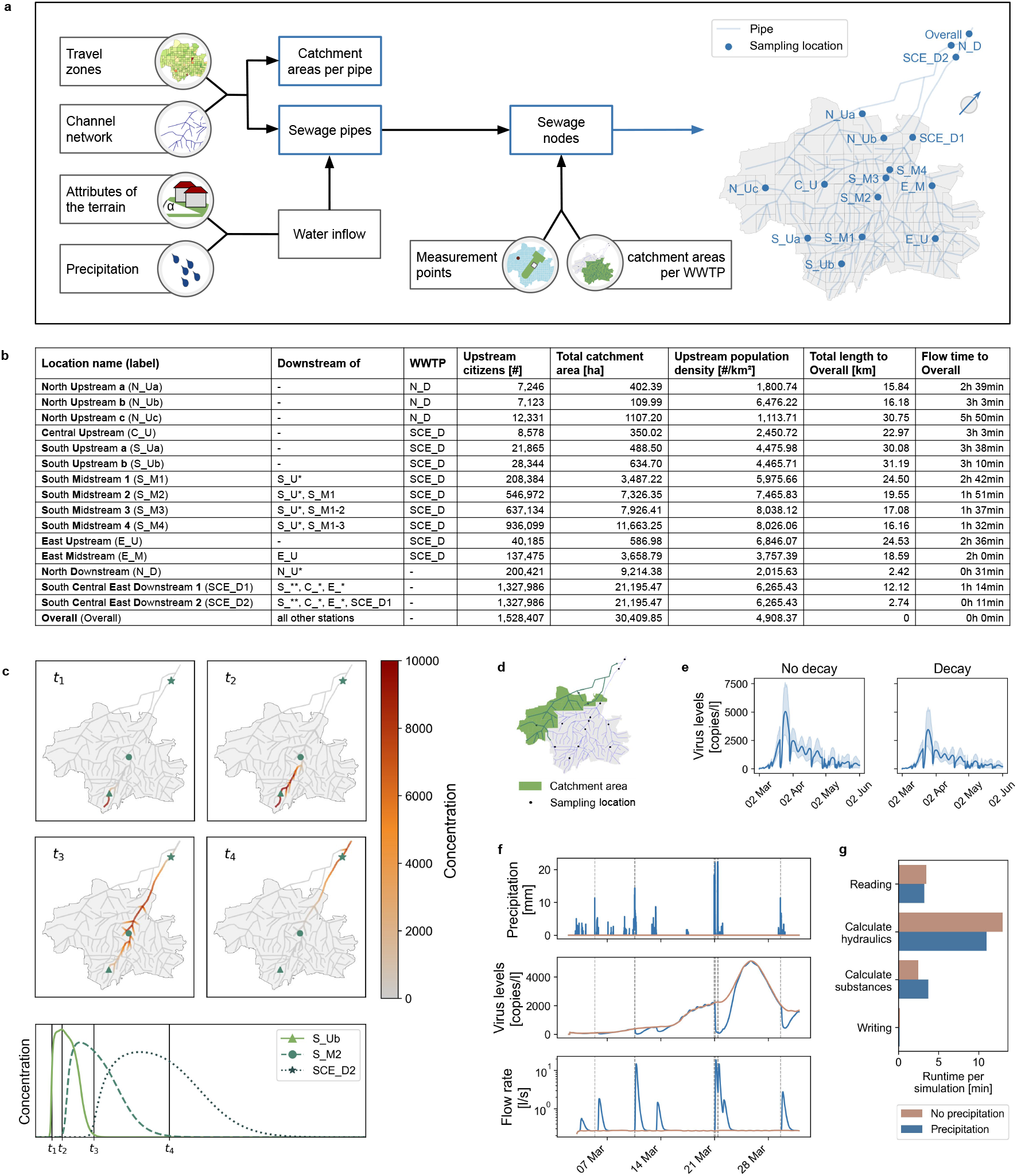
Overview of the wastewater model. **a)** Wastewater model definition pipeline. **b)** Characteristics of the sampling locations. **c)** Simulation of viral flow through the sewage from a point source: Throughout the simulation time frame, virus-free water from a small area in the south enters the sewage system. At simulation point *t*_0_, an additional source releasing water with a concentration of 10,000 copies/l is simulated. Over time, these virus copies travel downstream through the sewage, passing several sampling locations. **d)** Catchment area for location N_D. **e)** Influence of decay: Viral decay yields smaller virus levels at downstream location N_D. The shaded area indicates the 90% simulation interval over 100 simulations. **f)** Influence of precipitation at location N_D, for one example simulation: The dashed lines indicate a precipitation event *>* 10mm. **g)** Simulation runtime: One simulation takes about 20 minutes in total, and specific subsettings can influence runtime. Hardware details are provided in Supplementary Table S2.

The reconstruction followed a modular approach in which each component builds upon several data modalities and is validated against additional data sources. Initial area definitions were derived from traffic cells [31], which we further refined into 235 catchment areas to ensure that each principal pipe has at least one associated catchment area. The underlying sewer network, comprising approximately 2430 km of pipes within city boundaries, was reconstructed in a coarse-grained manner by georeferencing a high-resolution site plan from an official environmental report [40], thus yielding a network of around 340 km split into 318 pipe components. Terrain attributes like slope, degree of imperviousness, and population as well as precipitation events influence water inflows, which in turn determine pipe layout specifications, such as the diameter [41, 32]. A total of 305 nodes were defined not only by pipe junctions, but also by sewer sampling locations set up by researchers [42] and by retention basins [43, 40, 44]. Sewage in Munich generally flows toward the northeast, where two nodes correspond to the city’s wastewater treatment plants (WWTPs). Drainage plans were used to validate the catchment boundaries and flow directions [45, 46, 40]. We assumed a uniform rain distribution in Munich using measurement-based data from the area around Mariensäule, Munich, provided by the German Meteorological Service (DWD) [47] (Supplementary Fig. S4).

In the subsequent analysis, we focus on 16 sampling locations representing a diverse set of catchment area regions (North - N, South - S, Central - C, East - E) and sizes (Upstream - U, Midstream - M, Downstream - D) (Fig. 3b). In a test scenario decoupled from the ABM results, the simulation of inflow from a point source showcases that the model is capable of simulating how viral RNA travels through the sewage (Fig. 3c), reaching different sampling locations that each cover a specific catchment area (Fig. 3d, Supplementary Fig. S5). For the subsequent analysis, we take the 100 simulation outputs of the ABM described in the previous subsection and use them as input to the wastewater model. The model is evaluated under four scenarios: with and without viral decay, and with and without precipitation. Our results highlight how viral decay influences measurements, especially for downstream stations (Fig. 3e). On average over all simulations and time points, simulations with decay yield values of 8.3% smaller than simulations without decay at station N_Ua. In comparison, for downstream station N_D the difference is on average 30.8%. Precipitation events can lead to rainwater infiltration into the sewer system, which reduces virus levels while increasing flow rates (Fig. 3f). As our model accounts for groundwater storage and evaporation effects, the relationship between the amount of precipitation and increase of flow rates is non-linear. As this detailed modeling approach at the scale of a large city initially yielded simulation times of more than five hours per setting, we enhanced the simulation software by changing how inputs are loaded, reduced the amount of intermediate computations necessary for simulation, and changed how simulation outputs are stored. With these changes, simulations remain computationally feasible (Fig. 3g). We provide visualizations of the simulation results for all sampling locations and settings discussed in the following sections in the supplementary material (Supplementary Figs. S6 to S10).

### Calibration of transmission parameters reproduces observed epidemic trends

While most model parameters could be determined from the literature, the parameters controlling the transmission rates of infected agents were unknown. Thus, they were calibrated using Approximate Bayesian Computation (ABC) methods (Fig. 4a) against published data integrating reported case numbers with seroprevalence estimates (see Methods for details).

**Figure 4:**
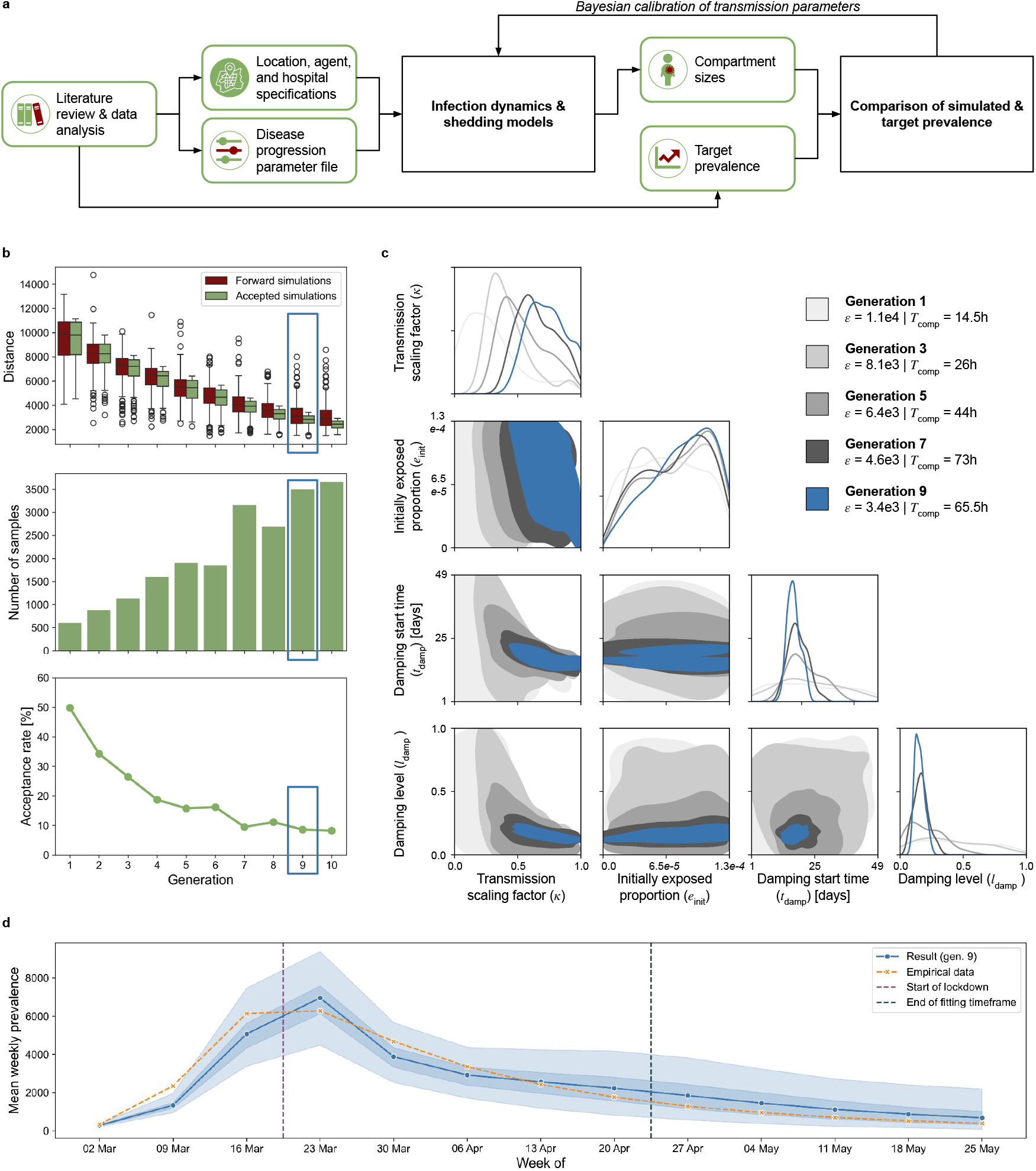
Model fitting results. **a)** Workflow for the Bayesian calibration of four unknown ABM parameters. **b)** Calibration diagnostics across generations: Euclidean distance between simulated and empirical prevalence for accepted simulations versus forward simulations with the same parameters; number of total samples; and acceptance rate. The generation used in this study is outlined in blue. **c)** Evolution of posterior parameter distributions, with the corresponding error values (*ϵ*) and computation times (*T*_comp_). As in (b), the final accepted generation is in blue. **d)** Agreement between simulated and empirical mean weekly prevalence; the period after the dark green dotted line was not included in the fitting process. The shaded blue areas indicate the 50% and 100% simulation intervals across the 300 accepted simulations from the final generation.

Due to the highly stochastic nature of the ABM, we monitored convergence of the ABC Sequential Monte Carlo algorithm over generations (Fig. 4b). By the ninth generation, improvements in the model fit were largely attributable to inherent model stochasticity rather than meaningful updates to parameter choices, as shown by the plateau in the median distance between simulated and observed prevalence for forward simulations with the selected parameters (top panel). This plateau, combined with the increasing number of total samples required to obtain 300 accepted simulations per generation (center panel) and the corresponding decline of the acceptance rate below ten percent (bottom panel), indicated convergence. Consequently, the calibration process was halted after the tenth generation, and the accepted simulations from generation nine, which showed stable results, were used throughout this study.

Each generation required up to about 80 hours of wall-clock time on a high-performance computing cluster (see Supplementary Table S2 for CPU details), with later generations generally taking longer due to increasingly strict distance thresholds and lower acceptance rates. Across all generations, approximately 21,000 simulations were executed, requiring a total of about 450 wall-clock hours. Posterior samples (Fig. 4c) indicate that the inferred transmission scaling factor (*κ*) and the damping parameters (*t*_damp_ and *l*_damp_) are tightly concentrated, suggesting good parameter identifiability, whereas the initially exposed proportion (*e*_init_) shows a broader distribution and exerted a weaker influence on model fit. The simulations selected via ABC exhibited good agreement with the observed data on the infection prevalence across the city, including the timing and magnitude of the epidemic peak (Fig. 4d).

### Temporal sampling design has minimal influence and virus normalization with pepper mild mottle virus can outperform normalization with flow rates

While simulations allow for very fine data resolution (we simulate four virus level measurements per hour), budget and personnel constraints make this infeasible in real-world contexts. We compared two commonly used temporal sampling protocols: a single morning grab and a 24-hour compound assembled from hourly grabs [6]. Using linear interpolation, we aligned both protocols to the simulated time series for direct comparison. The 24-hour compound is invariably influenced by changes in virus levels spanning at least one hour, whereas the daily grab is either unaffected or disproportionately affected (Fig. 5a). Consequently, differences between the daily grab and reference values are less frequent but more pronounced. This yields higher mean squared errors yet smaller mean absolute errors across simulations and time points (Fig. 5b). Thus, protocol choice materially alters inferred dynamics, even though overall errors are similar across protocols. This is consistent with the results for downstream sampling locations in our small-neighborhood study [27].

**Figure 5:**
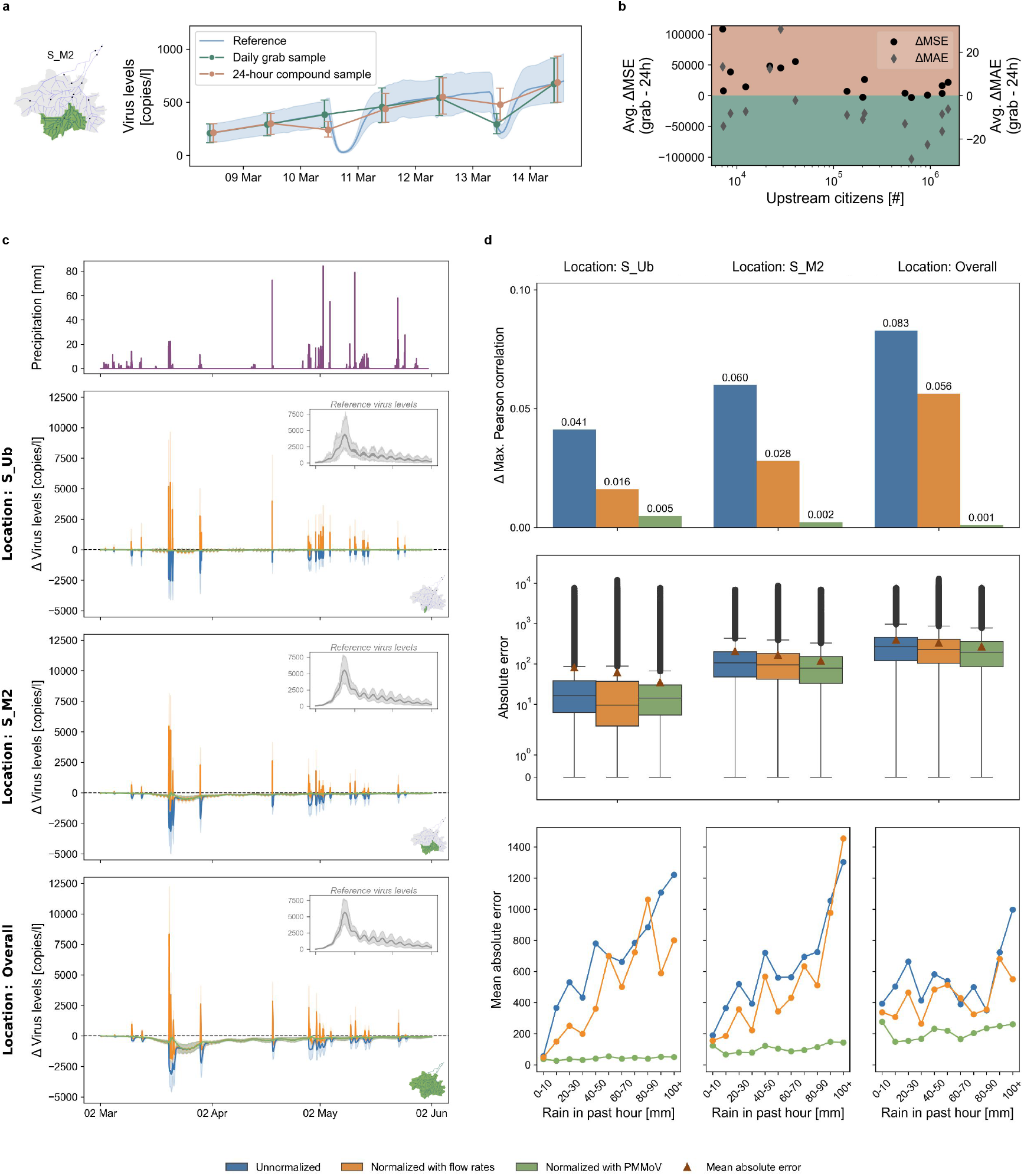
Effects of sampling protocol and normalization strategies. **a)** Simulated virus levels for 10:00am morning grab vs. 24-hour compound sampling (linear interpolation) with the reference (precipitation and decay). Shading shows the 90% simulation interval (100 simulations). **b)** Difference in Mean Squared Error (MSE) and Mean Absolute Error (MAE) to true virus levels across upstream citizens and simulations. Tan shading indicates that compound sampling yields better metrics than grab sampling, teal the opposite. **c)** Differences in virus levels between the rain- and decay-influenced scenario and the no-rain, no-decay reference scenario (in insets) under different normalization approaches. Shading shows the 90% simulation interval (100 simulations). **d)** Decrease in the maximal Pearson cross-correlation coefficient (averaged across simulations) between wastewater virus levels and prevalence relative to the reference scenario, by normalization approach and sampling location (top). Distribution of absolute errors across all simulations and time points by normalization and location (middle). MAEs across all simulations and time points against the past-hour rainfall, by normalization and location (bottom).

As precipitation events influence measurements (Fig. 3f), normalization of virus levels plays a key role in retrieving interpretable concentration values [48]. We analyzed normalization methods using additional measurements of either flow rates or the human fecal indicator pepper mild mottle virus (PMMoV) (see Methods for details). Our simulation results show that infiltration of rainwater to the sewer system causes greater proportional increases to the flow rates than decreases to the virus levels, yielding notable over-corrections when normalizing with flow rates (Fig. 5c). The further downstream the sampling location is in the network, the longer the average flow times in the corresponding catchment area. As a result, downstream measurements are more strongly affected by viral decay. Since neither of our normalization approaches can correct for viral decay, this leads to a systematically greater underestimation of the reference values from a no-precipitation and no-decay scenario at downstream locations. Normalization with PMMoV can recover reference values best and yields the largest maximum cross-correlation between virus levels and prevalence, with Pearson correlation coefficients as large as 0.93 (Fig. 5d). An increasing amount of precipitation significantly increases the error of the flow rate normalization strategy, which is not the case for the PMMoV-based approach.

### Virus levels can be used to infer prevalence distribution, while demographics can predict infection risk

We investigated how initial spatial heterogeneity in prevalence is preserved over time and whether it can be captured by wastewater surveillance. While holding the expected number of initially infected agents and all other parameters constant, we examined three spatial initialization regimes across wastewater catchments: Infections initially occurring all in one area in the city center, infections initially occurring only in one area at the city border, and a uniform initialization (default setting) (Fig. 6a). Using only five sampling locations, we observe substantially different virus level dynamics: Larger signals downstream of an initialized area help identify zones of higher prevalence, whereas similar magnitudes across locations are consistent with an approximately uniform distribution. Both local initialization scenarios produce an early concentration of cases in neighboring catchments. However, with central initialization the prevalence pattern approaches uniformity by the end of the simulation window. This convergence is plausibly driven by mobility between catchments: There is a net outflow from border areas (particularly in the south) and a relatively greater inflow to the center during working hours, yielding faster mixing over time (Fig. 2e).

**Figure 6:**
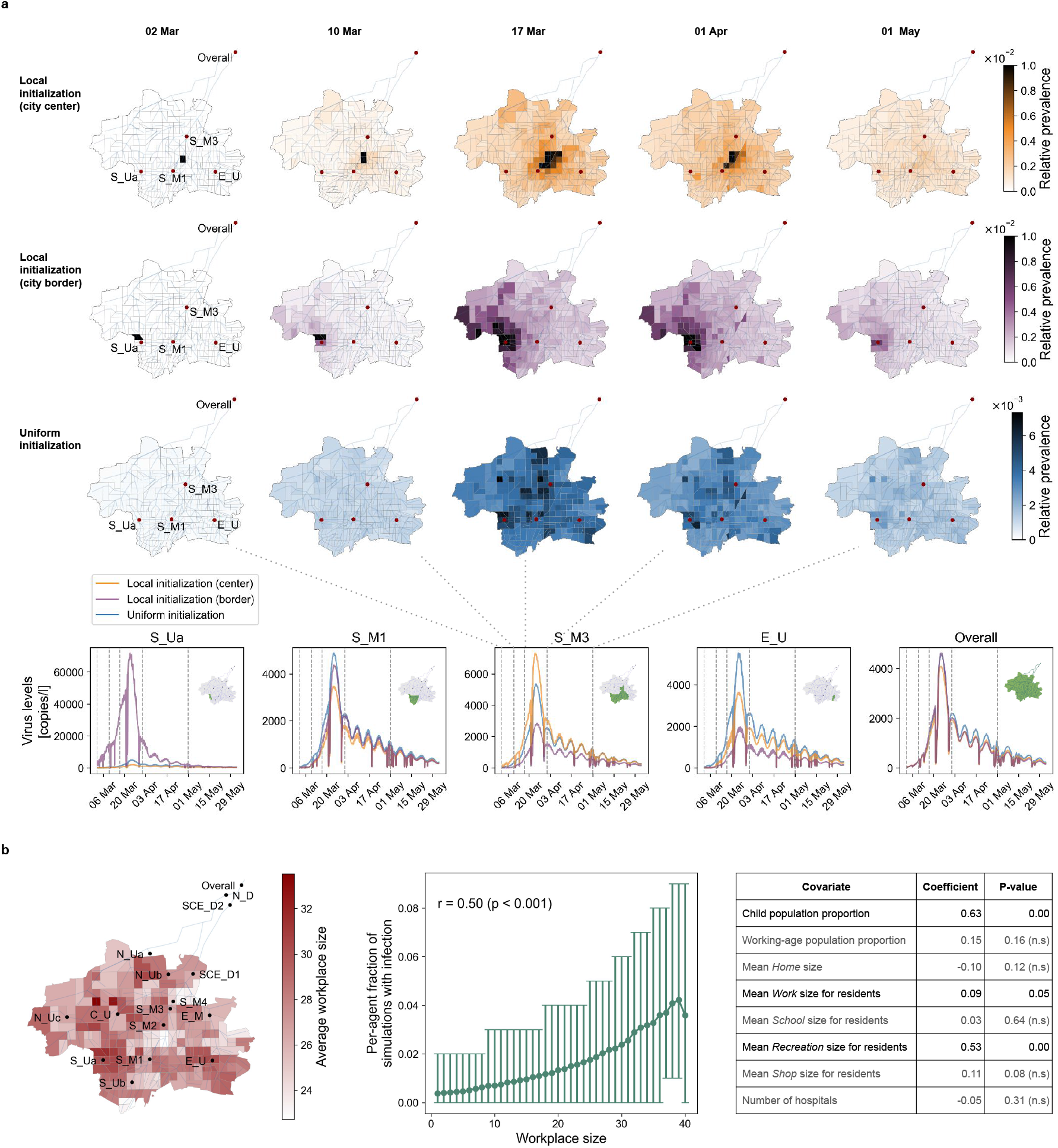
Factors influencing prevalence. **a)** Simulation outcomes for different initial prevalence distributions. Prevalence per area over time for local initialization between S_M1 and S_M3 (first row), local initialization directly upstream of S_Ua (second row), and the default uniform initialization (third row), with the corresponding average wastewater dynamics (bottom row). Shown are the averages over 100 simulations. **b)** Predictors of infection risk. Areas home to agents who commute to larger workplaces are concentrated in specific regions of the city (left). Working-age agents employed in larger workplaces tend to be infected more frequently across simulations (center); the error bars indicate the 90% simulation interval across 100 simulations. Ordinary least squares regression of area-level peak proportional prevalence (averaged across 100 simulations) on eight min-max normalized covariates indicates that areas with a high proportion of school-age residents, workers commuting to larger workplaces, and/or residents visiting larger recreation venues are more likely to become infection hotspots (right).

Even for a uniform initial prevalence distribution, some areas consistently show higher maximum prevalence values across 100 simulations (Fig. 6a). As the initialization of all agent attributes but the infection state was set constant across simulations, summary statistics like age distribution, location sizes, or number of hospitals can potentially influence area-level infection risk. Most transmissions occur at *Work* locations (Fig. 2b, Supplementary Fig. S3a), and relative prevalence varies across age groups (Fig. 2d). Our analysis yields larger child population proportion, mean *Work* size for residents, and mean *Recreation* size as key predictors of area-level infection risk (Fig. 6b).

### Transmission damping affects time lag between wastewater measurements and prevalence

To evaluate whether wastewater virus levels can serve as a reliable proxy for true infection prevalence, we analyzed the temporal cross-correlations between infection prevalence and RNA copies per liter measurable in wastewater at different sampling locations. Infection prevalence, summed respiratory transmission rates, summed RNA shedding rates, and the resultant wastewater virus levels are mechanistically interdependent (Fig. 7a), which underlies the potential of wastewater data to provide insights into disease transmission dynamics. There are strong cross-correlations between true total infection prevalence and summed respiratory transmission rates, with the latter time series leading the former by between three and four days (Fig. 7b); similarly, summed respiratory transmission rates lead cumulative 7-day incidence by between two and three days (Supplementary Fig. S11). Aside from minor variations in the magnitude of the time lag, this pattern is generally robust to changes of up to *±*20% in several key disease progression and shedding parameters (Fig. 7b), although such changes produce systematic shifts in the total number of transmissions, the maximum prevalence, and the timing of maximum prevalence (Fig. 7c).

**Figure 7:**
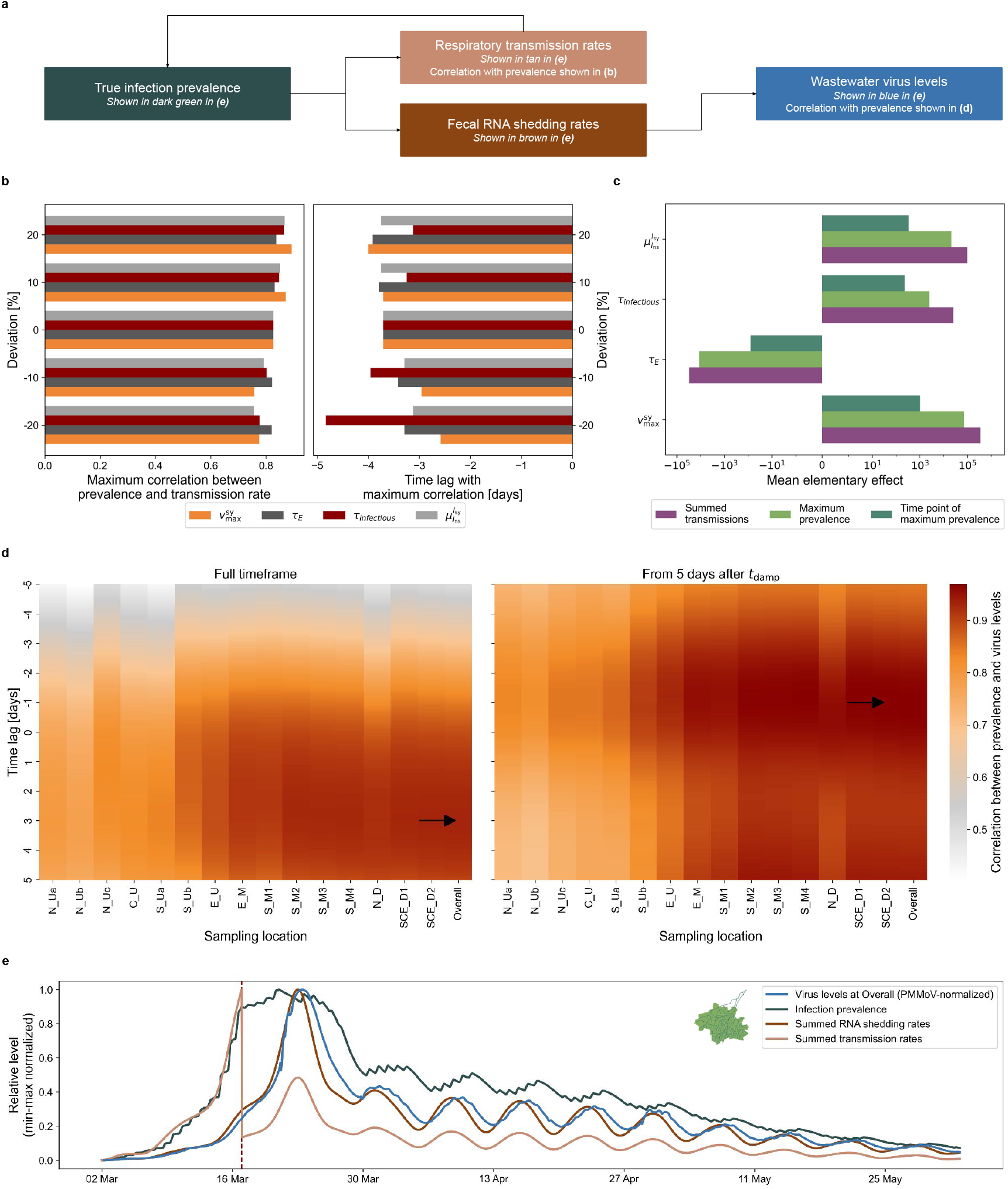
Cross-correlations between wastewater virus levels and prevalence. **a)** Schematic of causal links between prevalence, transmission, RNA shedding, and wastewater virus levels. **b)** Maximum cross-correlation between prevalence and summed transmission rates, averaged over 100 simulations, under changes in peak viral load 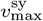, mean *Exposed* duration *τ*_*E*_, mean infectious period *τ*_infectious_, and symptomatic ratio 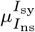 (left), with corresponding prevalence time lags (right). **c)** Elementary effects of these parameter changes on epidemic outcomes, averaged across 100 simulations per setting. Elementary effects are normalized differences from baseline. **d)** Cross-correlation coefficients between wastewater virus levels per location and total true prevalence, averaged across all 100 simulations, for two temporal windows. Time lags indicate prevalence shifts. Black arrows mark maxima. **e)** Trajectories of wastewater virus levels at the Overall sampling location, prevalence, shedding rates, and transmission rates for an example simulation in which transmission damping started on 17 March (red line).

Despite the dependence of viral decay on sewer travel time, the strongest cross-correlations between prevalence and wastewater virus levels occurred at sampling locations with larger catchment areas, particularly the southern midstream and downstream sites as well as the Overall location. The cross-correlation function reached its maximum when prevalence was shifted forward by approximately three days (Fig. 7d, left). This time-lag structure results from the transmission rate damping applied in mid-March to simulate the effects of NPIs: The damping reduced respiratory transmission rates but not fecal RNA shedding rates, causing a sudden drop in transmission potential and a plateau in prevalence while wastewater virus levels continued to climb (Fig. 7e). Consequently, in this context the prevalence peak precedes the wastewater virus level peak. When the analysis is restricted to the period after the damping effect stabilizes (Fig. 7d, right), downstream wastewater virus levels predict prevalence trends, since high wastewater virus levels indicate that the level of infectiousness across the population is also high and that a forthcoming rise in cases is therefore likely. These results demonstrate that public health interventions initiated at a single point in time can in the short term alter the temporal relationship between wastewater virus levels and infection prevalence, which must be considered when interpreting longitudinal wastewater surveillance data.

## Discussion

This study advances the coupled modeling of infection and wastewater processes by introducing a novel, city-scale framework that remains computationally tractable while reflecting conditions during the initial phase of the COVID-19 pandemic in Munich. Although access to detailed sewer system specifications may be restricted for security reasons, we show that it is feasible to construct a realistic hydrodynamic model by integrating heterogeneous publicly available data on population, infrastructure, and meteorology. The strong agreement between simulated and observed infection prevalence supports the validity of our calibration strategy and provides a solid foundation for subsequent analyses. By explicitly incorporating the effects of weather variability, sewer dynamics, viral decay, human mobility, viral shedding, and NPIs, our framework supports systematic evaluation of sampling protocols and normalization strategies. It also enables demonstrations of how appropriately analyzed wastewater data can be leveraged both to infer the spatial distribution of infections and to anticipate epidemic peaks shortly before they occur.

Our results on sampling and normalization strategies confirm the findings from our earlier small-scale study, illustrating their generalizability to realistically modeled cities. With respect to temporal sampling design, we find that 24-hour compound samples generally provide a more stable reflection of overall trends by averaging over short-term fluctuations. However, neither the compound nor the grab sampling approach consistently outperformed the other in our study, consistent with prior work reporting that both yield comparable results [49, 50]. As such, we recommend using normalization to adjust for dilution artifacts rather than trying to circumvent them via sample timing. In our simulations, flow-based normalization consistently over-corrected virus levels during precipitation-driven dilution events, which could lead to systematic overestimation of true prevalence. In contrast, normalization using the human fecal indicator PMMoV yielded more stable values, closely aligned with no-rain, no-decay reference scenario, and produced the strongest correlations with true infection prevalence. Our conclusions here are consistent with those of Isaksson et al. [51] but diverge from those of Rainey et al. [52] and Dhiyebi et al. [53], who found PMMoV normalization to be inferior to flow rate normalization when adjusting for population size changes and seasonality, respectively. While Rainey et al. did not account for rainfall, Dhiyebi et al. noted that infiltration can have counterintuitive effects on PMMoV concentrations, since increased flow can scour settled materials from the sewer walls. Our model did not incorporate this mechanism and may therefore overestimate the robustness of PMMoV normalization. Even so, our results highlight that flow rate normalization should only be implemented with caution in settings where rainwater infiltration is possible.

Our findings also demonstrate how NPIs can transiently alter the temporal relationship between wastewater virus levels and infection prevalence. Specifically, in our simulations, the transmission rate damping introduced in mid-March reversed the usual lag structure observed in our earlier study, where wastewater levels typically preceded prevalence by one to two days. This occurred because the damping, reflecting the effects of NPIs like mask mandates, reduced respiratory transmission and thereby slowed further increases in prevalence while fecal RNA shedding from existing infections continued unabated. Once the damping effects stabilized, the expected pattern of wastewater leading prevalence re-emerged. These findings highlight the need to interpret WBS data in the context of concurrent interventions and other population-level changes in transmission dynamics; they also contribute to ongoing discussion in the literature about time lags between wastewater data and reported cases. Many past studies have found that wastewater viral levels tend to precede reported case counts by between zero days and two weeks, reflecting both the feedback loop between shedding and prevalence as well as reporting delays inherent in clinical surveillance systems [54]. Observed offsets between wastewater data and cases may vary this widely across settings and epidemic phases for a variety of reasons, including testing capacity and biological differences between virus variants [55]. Our study suggests that public health interventions may also play a role in these discrepancies across studies, by decoupling prevalence and wastewater trends in the short term.

Even under uniform infection initialization, spatial variations in prevalence consistently emerged as a function of demographic and structural factors. In particular, areas with more children, residents commuting to larger workplaces, and attendance at larger recreation venues tended to have a higher infection risk. The distribution of recreation venue sizes was highly right-skewed, with only 62 out of nearly 37,000 venues having a capacity of more than 1,000 people, but this handful of very large locations helped drive higher prevalence in areas whose residents attended them, although only a small fraction of total transmission occurred at these locations before they were all closed in our model in mid-March. Thus, the correlation between recreation venue size and area-level prevalence highlights the disproportionate influence of large gatherings, consistent with the broader literature on the significance of super-spreading events [56, 57]. Similarly, workplace size distribution also emerged as a key determinant of infection risk at both the individual and area level, despite our model constraining workplace transmission by capping effective workplace sizes at 40 persons. In exploratory runs without this cap, virus levels at one upstream sampling location were dramatically elevated due to its position directly downstream of a 792-person workplace. While further refinement of workplace contact structures in our model is still needed, these findings reinforce that workplace size and organization are critical mediators of epidemic spread, as also evidenced in previous studies [58, 59, 60].

As spatial heterogeneity in infection prevalence emerges, our results indicate that WBS data can help identify areas of disproportionate risk. When infections were initialized locally rather than uniformly, differences in the magnitude of virus levels across catchments reflected meaningful information about local prevalence distributions during the transient phase before convergence to a citywide equilibrium. The eventual distribution of infections depended strongly on seeding location as well as commuting patterns and local area characteristics, underscoring the value of coupling wastewater-based epidemiology with demographic and mobility data to contextualize observed virus levels and inform targeted public health responses.

While our framework aims for realism, it necessarily depends on imperfect data, as all models do. In particular, empirical prevalence data for our setting of interest were substantially affected by underreporting, and although we therefore employed model-corrected estimates for parameter calibration, their unbiasedness cannot be assured. We also calibrated against total prevalence across Munich rather than catchment-specific prevalence, which was unavailable, limiting spatial accuracy. Additionally, empirical wastewater measurements in Munich were very sparse during the first wave of the COVID-19 pandemic and were collected only from mid-April 2020 onward, when the virus was only sporadically detectable in wastewater; those measurements that are available are not normalized and are subject to unquantifiable uncertainty. This limitation precluded formal Bayesian calibration of the wastewater dynamics sub-model. Collectively, these factors mean that the simulated spatial distribution of cases may not reflect reality, particularly during the early outbreak period. To mitigate this limitation, we explored wastewater surveillance effectiveness under multiple synthetic initialization scenarios.

Our modeling framework is designed to capture the complexity of viral shedding dynamics, and although we aimed to obtain empirical data to inform our representation of these processes, certain simplifying assumptions described in our previous work [27] have by necessity been retained. Specifically, due to a lack of available data on urinary and fecal shedding, we assume it to be proportional to respiratory shedding, with the constant of proportionality changing when transmission rate damping is applied. We further assume continuous shedding into the wastewater system, despite the likelihood that domestic wastewater production follows a circadian rhythm, with higher output in the morning and evening and reductions at night. Relaxing these assumptions to explicitly incorporate additional sources of shedding variability into our model could increase short-term fluctuations in simulated wastewater virus levels. This might, in turn, increase the advantages of 24-hour compound sampling over grab sampling and reduce cross-correlations between prevalence and wastewater measurements during low-prevalence periods.

Given the importance of socioeconomic heterogeneity in shaping transmission dynamics, we explicitly consider it in our modeling where possible, though its representation is unavoidably limited by the availability of suitable data. Differences in household and workplace size distributions across city sub-regions, as captured by traffic data, are reflected in our model, but our representation of NPIs is simplified. For example, we represented work-from-home orders by randomly closing 30% of workplaces citywide, when in reality, remote work is more feasible among individuals with higher-income office-based occupations whose homes may be geographically clustered [38], influencing the spatial heterogeneity of transmission. We also did not model the gradual reopening of stores starting from mid-April 2020 or schools from mid-May, resulting in modeled mobility reductions that are both uniform across the city and likely over- or underestimated at specific time points. Refining the model to fully capture the interactions between socioeconomic factors, mobility changes, and the spatial distribution of cases would enable evaluation of how targeted wastewater sampling could provide meaningful insights into how the burden of disease varies by subpopulation.

In summary, our study establishes the feasibility of a city-scale integrative framework that links agent-based modeling of infection dynamics with hydrodynamic modeling of wastewater processes, moving beyond the constraints of previous approaches to offer new mechanistic insights into how wastewater signals relate to underlying infection dynamics. We show that normalization using human fecal indicators can enhance the robustness of wastewater surveillance and identify demographic and structural features that drive local heterogeneity in infection risk. Crucially, we demonstrate how NPIs can temporarily reshape the time-lag structure between wastewater signals and infection prevalence. Together, these results support the integration of wastewater surveillance data into predictive modeling pipelines, not only for COVID-19 but also for future emerging pathogens.

## Methods

### Model Overview

Our integrative modeling framework consists of coupled state-of-the-art models for the spread of infectious diseases and wastewater dynamics linked via a viral shedding model. The combined framework has a modular structure, meaning that individual components can be updated or replaced without the rest of the framework being affected.

The *infection dynamics model* is an ABM that describes both the location and the infection state of individuals (agents) over time, with the latter subject to change due to disease transmission, development or worsening of symptoms, recovery, or death.

The *shedding model* describes the time-dependent rates at which infected agents release virus particles into their surroundings. This model uses the agent infection histories from the infection dynamics model as inputs. In turn, it informs (a) the probability of respiratory transmission from infected to co-located susceptible agents in the infection dynamics model and (b) the amount of viral RNA released via urine and stool into the sewer system of the wastewater model. Both respiratory and fecal/urinary shedding are dependent on the agent’s viral load, which increases after infection before declining due to the host’s immune response.

The *wastewater model* describes the sewage flow and transport of viral RNA within the sewer network of the region of interest. This model uses as inputs the fecal/urinary shedding levels from the viral shedding model and the agent locations from the infection dynamics model. In turn, it calculates the RNA concentrations over time and across the sewer network, accounting for the inflow of domestic wastewater as well as the infiltration of storm- and groundwater, viral decay, the architecture of the sewer system, and the slope of the terrain.

### Infection dynamics model

To define our infection dynamics model we enhance our previously proposed ABM [27] by incorporating NPIs, integrating real-world data, and optimizing Input/Output routines. This ABM is implemented in the C++-based software MEmilio [61] and models individuals, called agents, with different attributes that can be either static (e.g., age group and assigned locations) or dynamic (e.g., current location and infection state) (Fig. 2a). Throughout the simulation time frame, agents move between locations of different types according to mobility rules. The considered location types for this study are *Home, School, Work, Recreation, Shop, Hospital*, and *Intensive Care Unit (ICU)*. There can be multiple locations of the same type in the model, but every agent is assigned at most one location per type.

Infectious agents can transmit the virus to susceptible agents at the same location. For a susceptible agent, the waiting time until potential transmission is assumed to be exponentially distributed with the rate parameter corresponding to the cumulative transmission rate of all infectious agents present at their current location. Upon exposure, an agent’s infection trajectory, including the agent-dependent subsequent infection states and corresponding transition time points, is sampled. Infection duration differs across agents due to both the stochastic nature of infection state stay times and the possibility of experiencing no, mild, severe, or critical symptoms. The infection states used for this study are *Susceptible* (*S*), *Exposed* (*E*), *Non-symptomatically Infected* (*I*_ns_), *Symptomatically Infected* (*I*_sy_), *Severely Infected* (*I*_sev_), *Critically Infected* (*I*_cri_), *Recovered* (*R*), and *Dead* (*D*).

The ABM for Munich provides the possibility of realizing NPIs via location closures or transmission rate dampings. *Transmission rate dampings* modulate the infectiousness of agents over time and are defined by the time point *t*_damp_ at which the damping becomes active and the corresponding damping level *l*_damp_. The damping level is a multiplicative factor applied to the transmission scaling parameter *κ*_*λ*_, thus reducing the transmission rate of all infectious agents, from time point *t*_damp_ onward until either the damping is lifted or replaced by a new one.

While transmission rate dampings apply globally and can be used to model NPIs like social distancing or mask wearing that do not target specific location types or age groups, measures like school closures, home office regulations, or reduced availability of recreation facilities can be realized via *location closures*. A location closure (*τ*_closure_, *t*_closure_, *l*_closure_) randomly closes a proportion *l*_closure_ of all locations of type *τ*_closure_ ∈ {*School, Work, Recreation, Shop*} from time point *t*_closure_ onward. The closure remains active until either locations are reopened or until it is replaced by another location closure affecting the same type.

As described in the Results section, the DEMO data set used to initialize the ABM only contains 1,312,903 persons. To scale up to the full population size of 1,536,984, we created 224,081 new agents by sampling their properties from the underlying data set. For each new agent, household size and home traffic cell were sampled first and a new *Home* location was then created within the sampled home traffic cell for each new household. Age was sampled next, conditional on the household size since age distributions differ between households of different sizes (Supplementary Fig. S1a-c). Finally, the new agents were assigned to existing locations of each non-*Home* type.

### Shedding and transmission model

For this study we utilized our previously published shedding model [27], which is briefly summarized below. Infectious individuals transmit the virus via droplets and aerosols to other individuals and shed viral RNA into the sewage system. We model an agent’s respiratory and fecal shedding over time as proportional to a sigmoidal function of its viral load:

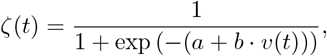

where *a, b >* 0 are shape parameters and *v*(*t*) the individual viral load [62, 63]. The agent-dependent viral load increases from the time point of exposure, then peaks and decreases as the host’s immune response takes effect until it reaches zero again at the time point of recovery or death (see Supplementary Eq. (1)). We assume that the RNA shedding rate and the transmission rate remain negligible during the *Exposed* phase, which is modeled by a time shift in the sigmoidal curve *ζ* [27]. The sigmoidal function *ζ*(*t*) is translated into an RNA shedding rate and a transmission rate by multiplication with scaling factors *κ*_*γ*_ and *κ*_*λ*_, respectively.

### Wastewater model

The wastewater network is formalized as a directed-acyclic graph whose edges represent pipes and whose nodes represent manholes and special constructions like retention basins. Each edge is parameterized by a corresponding length, diameter, elevation, and resultant maximal storage volume, as well as a catchment area. The catchment area defines the water inflow in the form of domestic and rain inflow. The instantaneous system state comprises water level as well as flow rates. Each node is parametrized by its storage volume, describing discharge and solute concentrations. The presented Munich model is an approximation combining open-source real-world information on measurement points, retention basins, the slope of the terrain, channel network, and catchment areas with the presented traffic zone data and domain knowledge to define plausible nodes, edges, and areas. Measurement stations can be defined at any manhole of the wastewater network. External inflow to the sewage is driven by two boundary processes: Precipitation-induced surface runoff and anthropogenic discharge from domestic water use, so the model can reproduce both precipitation-event hydraulics and dry-weather base flow. Unsteady pipe flow is described by the fully dynamic 1D Saint-Venant equations discretized in space along each pipe segment and in time. Mass and momentum conservation at each node close the system, while energy losses from pipe friction (Prandtl-Colebrook equation) and minor losses at manholes (Borda-Carnot equation) refine the simulation. Using an Euler scheme to solve the equation, water levels are iteratively updated based on discharges until the end of the simulation time frame. Once the time-resolved hydraulic fields are known, advective–dispersive transport and decay of dissolved constituents are superimposed to obtain spatiotemporal concentration profiles. Viral genomes released by agent-based shedding events enter the network and are conveyed downstream according to the computed flow field, allowing prediction of virus levels at any sampling location. We assume wastewater production to be proportional to the number of agents at a specific location. In this two-stage workflow (hydrodynamics first, reactive transport second) the model yields high-fidelity, geospatially explicit estimates of virus levels in the wastewater network. For more details on the simulation procedure, we refer to the description in our previous work [27].

We assumed exponential decay of RNA in wastewater with a 90-percent reduction time of 5.7 days or 492,480 seconds, based on an estimate for the 90-percent reduction time of the E biomarker of endogenous SARS-CoV-2 RNA in aqueous wastewater samples from the Luxembourg City WWTP at 12 ^°^C [64]. As such, in our model RNA(*t*) = RNA(0) · exp(−*kt*), with RNA(*t*) denoting the virus levels in copies per liter at *t* seconds and 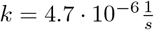.

During precipitation events, rainwater can infiltrate sewers, causing PMMoV concentrations to be diluted and flow rates to rise. Hence, adjusting results by the expected-to-measured PMMoV concentration ratio or the measured-to-expected flow ratio can help curb unwanted variability. Normalization of virus levels is calculated according to one of the two formulas:

- normalization with flow rates: 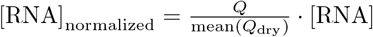
- normalization with PMMoV: 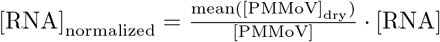

where *Q* is the flow rate, mean(*Q*_dry_) the mean flow rate on dry days, [PMMoV] the PMMoV concentration, and mean([PMMoV]_dry_) the mean concentration of PMMoV on dry days. As in our previous work [27], we simulated PMMoV concentrations by assuming a constant per-agent shedding over time and negligible substance decay.

We evaluated two sampling strategies: The first consisted of a single grab sample taken at 10:00am each day, and the second was a 24-hour compound sample, defined as the mean of 24 hourly samples per day with a nominal reporting time of 12:00pm. For both strategies, the resulting daily values were linearly interpolated in order to enable comparison with the high-resolution simulation outputs.

Wastewater model settings used in this study are summarized in Supplementary Table S3. The model definition and simulations were implemented using the software ++SYSTEMS [39].

### Model parameterization and calibration

For this study, our simulation time frame consisted of a 92-day period from 12am on Monday, 02 March 2020 to 12am on Tuesday, 02 June 2020. During this period, only the wild-type SARS-CoV-2 variant was in circulation [65]. Thus, values for all disease progression parameters [66] and all shedding parameters (except for the transmission scaling parameter *κ*_*λ*_) [62, 63] were taken from the literature to reflect wild-type COVID-19 (see Supplementary Table S4). Except for the number of initially exposed persons, initial conditions (corresponding to the state at 12am on 02 March 2020) were set such that on average, 75 agents started in state *I*_ns_, 30 in state *I*_sy_, and 0 across states *I*_sev_, *I*_cri_, *R*, and *D* [67]. Additionally, to account for NPIs not related to location closures, such as mask-wearing and social distancing, a single transmission rate damping was applied.

The transmission scaling factor *κ*_*λ*_, the initially exposed proportion *e*_init_, the transmission damping level *l*_damp_, and the damping start time *t*_damp_ could not be drawn with confidence from the literature and were therefore used as the four free parameters in the calibration process for our ABM. Broad, uninformative priors were used for *κ*_*λ*_, *l*_damp_, and *t*_damp_. We assumed that the number of agents in the *Exposed* state at the beginning of March 2020 should not exceed approximately twice the number of agents in the infectious states *I*_ns_ and *I*_sy_ due to the relatively short incubation period of wild-type SARS-CoV-2 [66]. Accordingly, to ensure that no more than about 200 agents out of the approximately 1.54 million persons in Munich started in the *Exposed* state, the prior distribution for *e*_init_ was set to *U* (0, 0.00013).

Calibration of the ABM was performed using Approximate Bayesian Computation methods with sequential Monte Carlo sampling via the python package pyABC [68]. As a calibration target, we utilized a previously published estimate of SARS-CoV-2 prevalence in Munich over time from the beginning of March to late April 2020, based on the integration of reported case and seroprevalence data [67]. Specifically, the Euclidean norm was minimized between the weekly averages of this estimate of the true prevalence and the corresponding weekly averages of our model’s total simulated prevalence across the *I*_ns_, *I*_sy_, *I*_sev_, and *I*_cri_ states. The calibration process was terminated once further improvements in the model fit between successive generations were attributable primarily to stochastic variation rather than systematic changes in parameter values, indicating convergence. Of the 300 accepted simulations in the final generation for which parameter-related fitting improvements were seen, 100 were randomly selected for use in the simulations for this study.

As virus levels scale linearly with changes to the shedding magnitude, we estimated the shedding scaling factor *κ*_*γ*_ post-hoc given simulations with an initial shedding magnitude *k*_0_. Assuming a log-normal noise distribution, the estimate 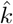 was obtained by applying the optimizer L-BFGS on real-world measurements of virus levels above the detection threshold from across Munich in April and May 2020 [42]. As a result, the simulated virus levels are in the expected range and trend downward as the empirical prevalence did during the period of interest (Supplementary Fig. S12).

## Author contributions

J.B.: conceptualization, methodology, software, formal analysis, investigation, writing – original draft, writing – review & editing, visualization; N.T.: conceptualization, methodology, software, formal analysis, investigation, writing – original draft, writing – review & editing; K.W.S.: conceptualization, methodology, formal analysis, investigation, writing – original draft, writing – review & editing, visualization; N.S.: conceptualization, methodology, formal analysis, investigation, writing – original draft, writing – review & editing, visualization; A.F.H.: methodology, software, writing – review & editing, funding acquisition; S.K.: methodology, software, writing – review & editing; A.S.: methodology, data curation, writing – review & editing; J.J.: methodology, data curation, writing – review & editing; A.W.: validation, writing – review & editing, supervision, funding acquisition; M.J.K.: validation, writing – review & editing, supervision, funding acquisition; J.H.: validation, writing – review & editing, supervision, project administration, funding acquisition.

## Implementation and availability

The version of MEmilio used in this study is publicly available at https://github.com/SciCompMod/memilio/tree/inside-demonstrator-munich. MEmilio input files are provided either directly on GitHub (for small files) or on Zenodo (for large files) at https://doi.org/10.5281/zenodo.17096975. (The Zenodo record will be published once the peer review process is complete.) Access to the software ++SYSTEMS is available to researchers for scientific purposes upon request to; all ++SYSTEMS model files and simulation outputs used in this study are archived on Zenodo. Results were analyzed using the custom Python code available at https://github.com/schminin/INSIDeMunich.

