## Supplementary Material for "Coupled Epidemiological and Wastewater Modeling at the Urban Scale: A Case Study for Munich"

* Shared first author in alphabetical order

### A Supplementary transmission model

Here, we provide the function for an agent's viral load, denoted as $v(t)$, which is used to calculate their respiratory and fecal shedding over time. On the $\log_{10}$ scale, $v(t)$ is given by

$$v(t) = \begin{cases} \dfrac{v_{\max}^{\mathrm{sy}}}{\tau_E + \tau_{I_{\mathrm{ns}}}} \cdot (t - t_E), & \text{if } t \in [t_E, t_{v_{\max}}] \\ v_{\max} + -\dfrac{v_{\max}}{t_{\mathrm{R/D}} - t_{v_{\max}}} \cdot (t - t_{v_{\max}}), & \text{if } t \in (t_{v_{\max}}, t_{\mathrm{R/D}}] \\ 0, & \text{otherwise} \end{cases} \tag{1}$$

with $\tau_E, \tau_{I_{ns}}$ denoting the time (in days) the agent spends in the *Exposed* and *Non-Symptomatically Infected* states, respectively, and $t_E$, $t_{v_{\max}}$, $t_{\mathrm{R/D}}$ denoting the time points (in days) of virus exposure, maximal viral load, and recovery or death, respectively [1, 2, 3].

### B Supplementary tables

| Type | Number |
|---|---|
| DEMO trips | 4,750,458 |
| Agents obtained from DEMO trips | 1,312,903 |
| All ABM agents | 1,536,984 |
| All locations | 1,043,855 |
| *Home* locations | 922,604 |
| *School* locations | 3,422 |
| *Work* locations | 63,674 |
| *Recreation* locations | 36,924 |
| *Shop* locations | 17,184 |
| *Hospital* locations | 30 |
| *ICU* locations | 17 |
| Locations in catchment area | 996,369 |

**Supplementary Table S1: ABM Munich initialization details.**

| Task | Cores [#] | CPU specifics |
|---|---|---|
| Wastewater simulations | 8 | AMD Ryzen 9 5900HX |
| ABM scaling test | 14 | Intel Xeon “Skylake” Gold 6132 |
| ABM fitting & simulations | 48 | 2x AMD EPYC 7F72 3.20 GHz / 7443 2.85 GHz |

**Supplementary Table S2: Hardware specifics per computation task.**

| ID | MEmilio Initialization | Rain Scenario | Viral Degradation | Post-Processing |
|---|---|---|---|---|
| 1 | Uniform | No precipitation | No | None |
| 2 | Uniform | Precipitation | No | None |
| 3a | Uniform | Precipitation | Yes | None |
| 3b | Uniform | Precipitation | Yes | 24-hour compound sampling |
| 3c | Uniform | Precipitation | Yes | Daily grab sampling |
| 3d | Uniform | Precipitation | Yes | PMMoV normalization |
| 3e | Uniform | Precipitation | Yes | Flow rate normalization |
| 4 | Local (city center) | Precipitation | Yes | None |
| 5 | Local (city border) | Precipitation | Yes | None |

**Supplementary Table S3: Overview of wastewater model scenarios in this study.**

| Parameter | Value (per age group) |
|---|---|
| $\mu_{I_{\mathrm{ns}}}^{I_{\mathrm{sy}}}$ | 0.75 (0-15), 0.8 (16-80+) |
| $\mu_{I_{\mathrm{sy}}}^{I_{\mathrm{sev}}}$ | 0.0075 (0-15), 0.019 (16-34), 0.0615 (35-59), 0.165 (60-79), 0.225 (80+) |
| $\mu_{I_{\mathrm{sev}}}^{I_{\mathrm{cri}}}$ | 0.075 (0-34), 0.15 (35-59), 0.3 (60-79), 0.4 (80+) |
| $\mu_{I_{\mathrm{cri}}}^{D}$ | 0.05 (0-15), 0.14 (16-59), 0.4 (60-79), 0.6 (80+) |
| $\tau_E$ | mean: 3.335 (All); variance: 0.1189 (All) |
| $\tau_{I_{\mathrm{ns}}}^{I_{\mathrm{sy}}}$ | mean: 1.865 (All); variance: 0.1182 (All) |
| $\tau_{I_{\mathrm{ns}}}^{R}$ | mean: 7 (All); variance: 0.4999 (All) |
| $\tau_{I_{\mathrm{sy}}}^{I_{\mathrm{sev}}}$ | mean: 10.5 (0-34), 6 (35-80+); variance: 0.5954 (0-34), 0.2662 (35-80+) |
| $\tau_{I_{\mathrm{sy}}}^{R}$ | mean: 5 (0-15), 6 (16-34), 8 (35-59), 10 (60-79), 15 (80+); variance: 0.2689 (0-15), 0.2662 (16-34), 0.2636 (35-59), 0.2624 (60-79), 1.0562 (80+) |
| $\tau_{I_{\mathrm{sev}}}^{I_{\mathrm{cri}}}$ | mean: 5 (All); variance: 1.1957 (All) |
| $\tau_{I_{\mathrm{sev}}}^{R}$ | mean: 7 (0-34), 17.5 (35-79), 12.5 (80+); variance: 1.1142 (0-34), 3.2941 (35-79), 1.6807 (80+) |
| $\tau_{I_{\mathrm{cri}}}^{D}$ | mean: 6 (0-34), 16.5 (35-79), 11 (80+); variance: 1.1434 (0-34), 0.5896 (35-79), 0.262 (80+) |

**Supplementary Table S4: ABM transmission parameters.**

### C Supplementary figures

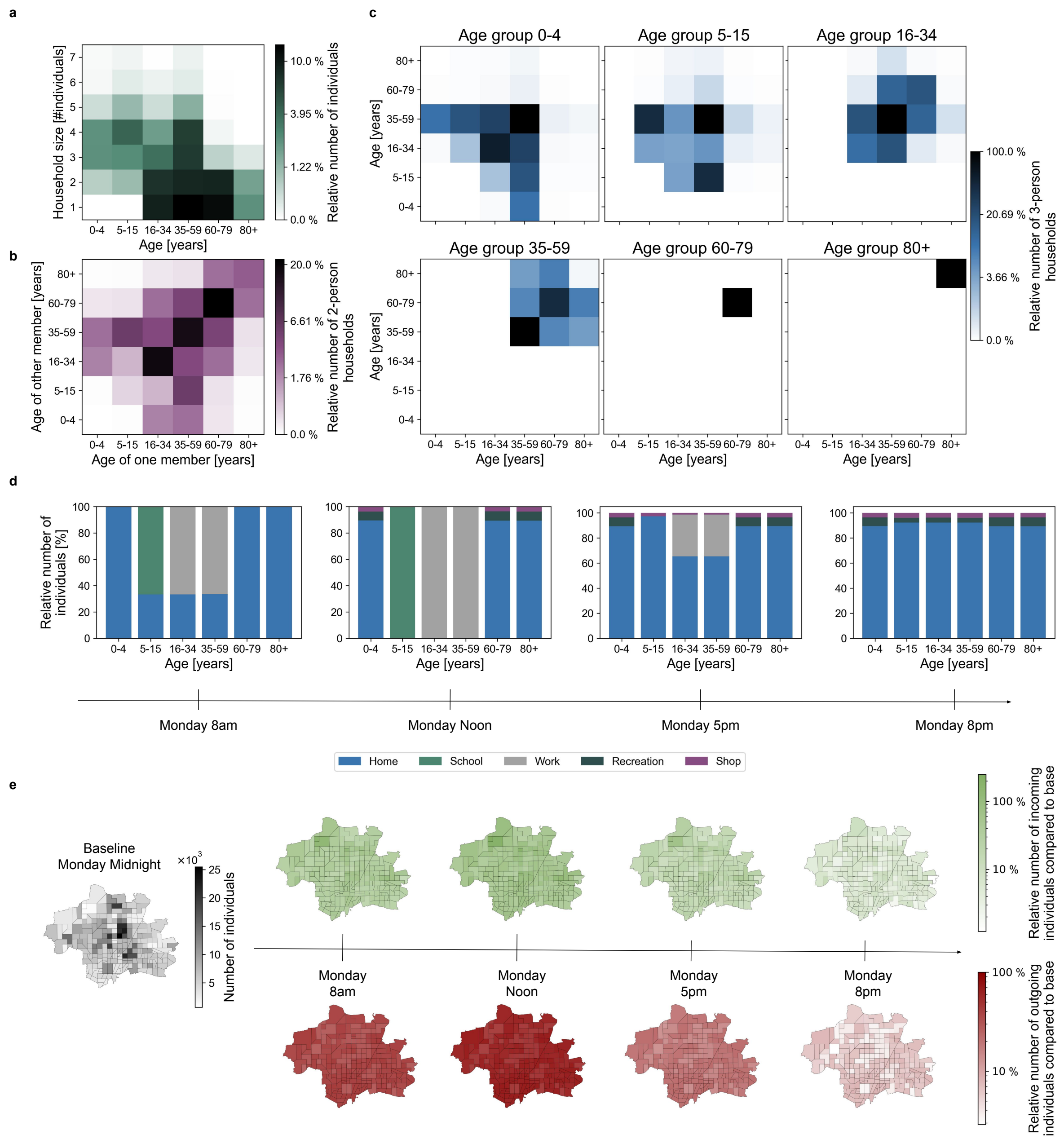


**Supplementary Figure S1: ABM details. a)** Age structure of the modeled population across household sizes. **b)** Age distribution of 2-person households. **c)** Age distribution of 3-person households conditional on the age of the youngest household member. **d)** Evolution of location type distributions during the day resolved by age group. Age group 5-15 is school-age while age groups 16-34 and 35-59 are working-age. **e)** Movement of agents between wastewater areas throughout the day. Shown are the relative differences in the number of agents, separated by incoming (non-residents) and outgoing agents (residents), compared to the baseline of Monday at midnight, when all agents are at *Home*.

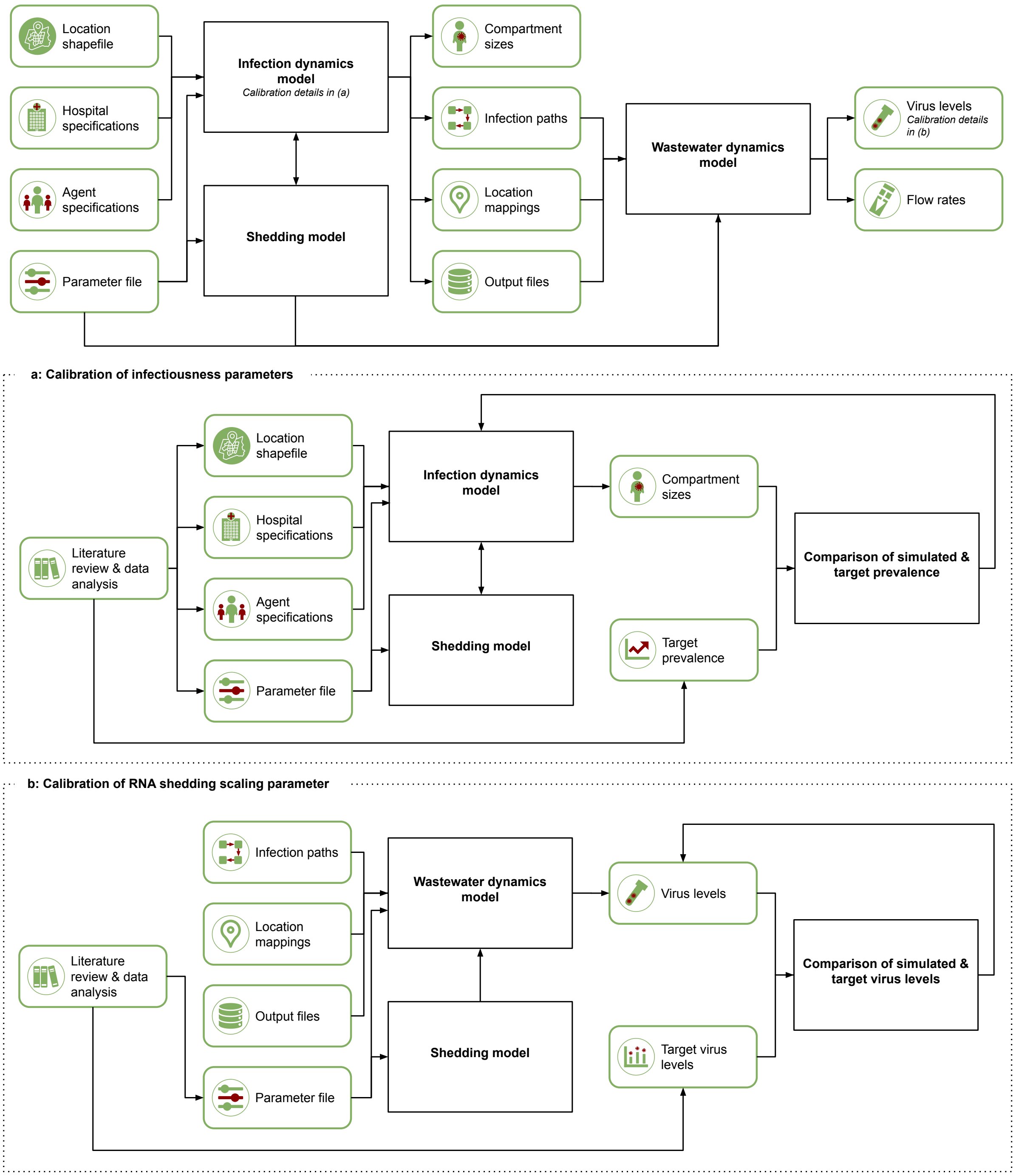


**Supplementary Figure S2: Model workflow.** The flow of information from user inputs to the model components and ultimately to the final outputs. **a)** Sub-workflow for calibrating the unknown infectiousness parameters within the infection dynamics model: the transmission scaling parameter $\kappa_\lambda$, the initially exposed proportion $e_{\text{init}}$, the transmission damping level $l_{\text{damp}}$, and the damping start time $t_{\text{damp}}$. **b)** Sub-workflow for calibrating the RNA shedding scaling parameter $\kappa_\gamma$.

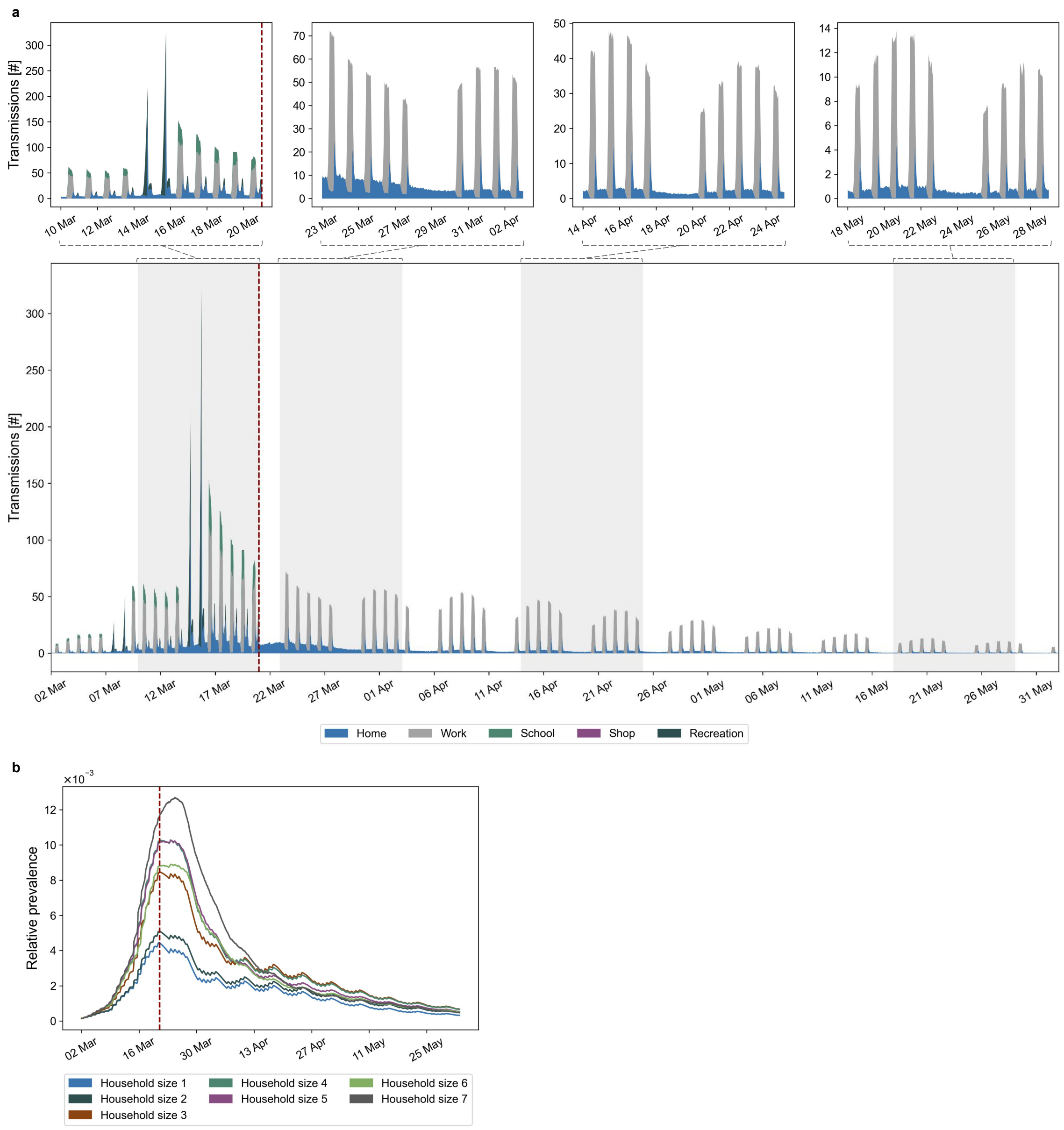


**Supplementary Figure S3: Prevalence and transmission dynamics over time. a)** Mean number of transmissions over time for each location type, across 100 simulations. The red line indicates day 19 of the simulated time frame, when location closures begin. **b)** Relative prevalence over time for every household size. Shown are the mean values across 100 simulations.

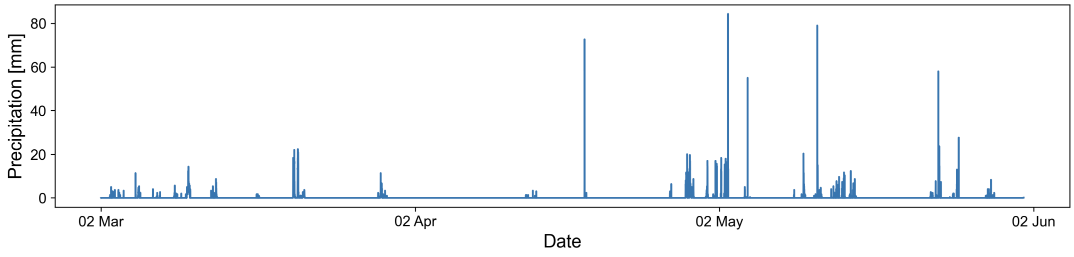


**Supplementary Figure S4: Precipitation over time.**

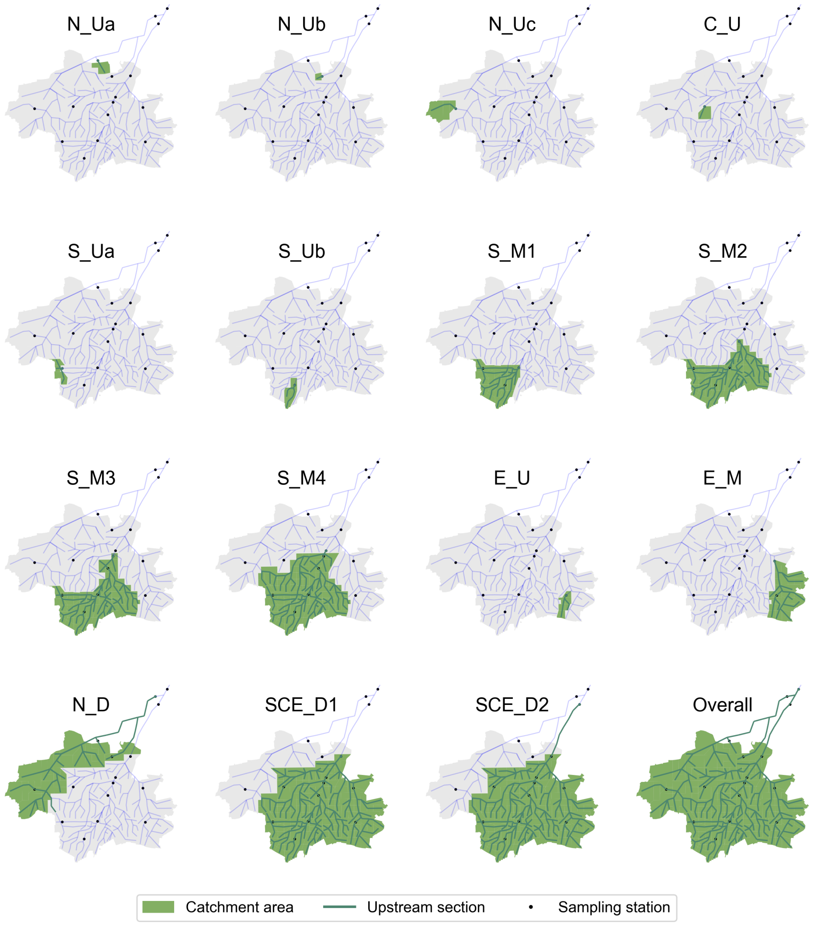


**Supplementary Figure S5: Catchment areas per sampling location.**

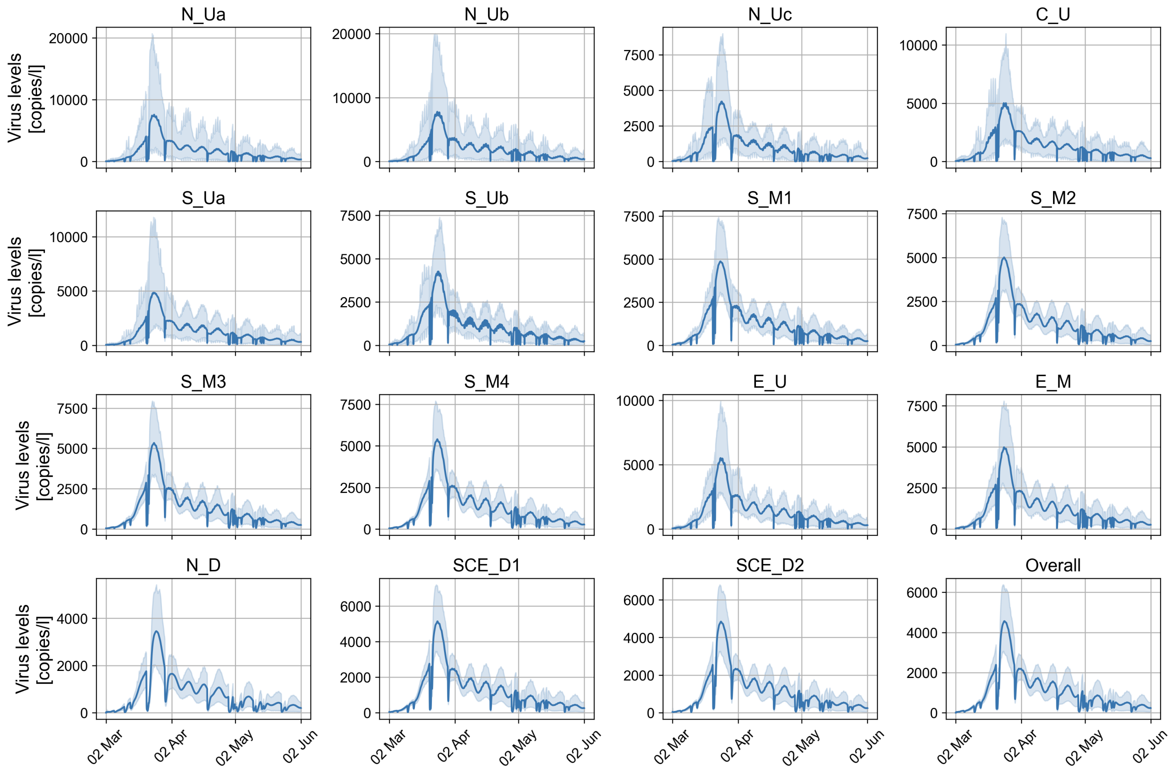


**Supplementary Figure S6: Virus levels per sampling location for the decay & rain scenario (uniform initialization of prevalence).** This corresponds to Scenario 3a in Supplementary Table S3 and reflects the standard experimental setup.

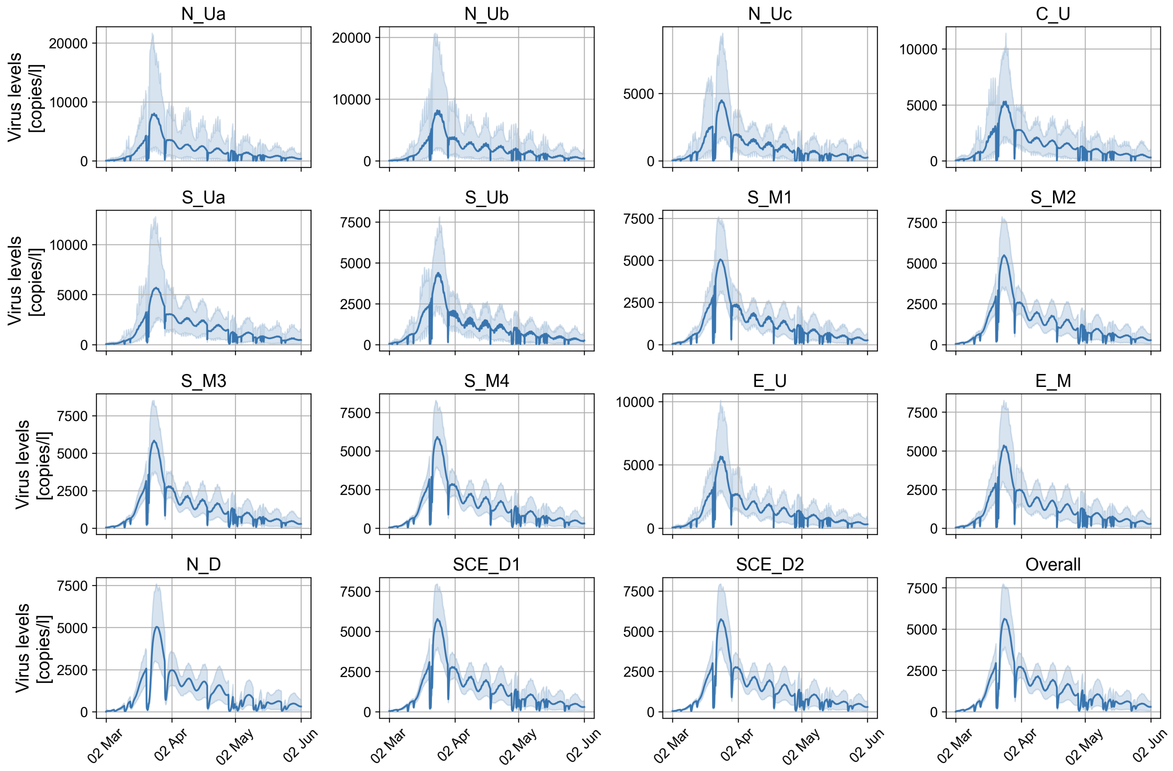


**Supplementary Figure S7: Virus levels per sampling location for the no decay & rain scenario (uniform initialization of prevalence).** This corresponds to Scenario 2 in Supplementary Table S3.

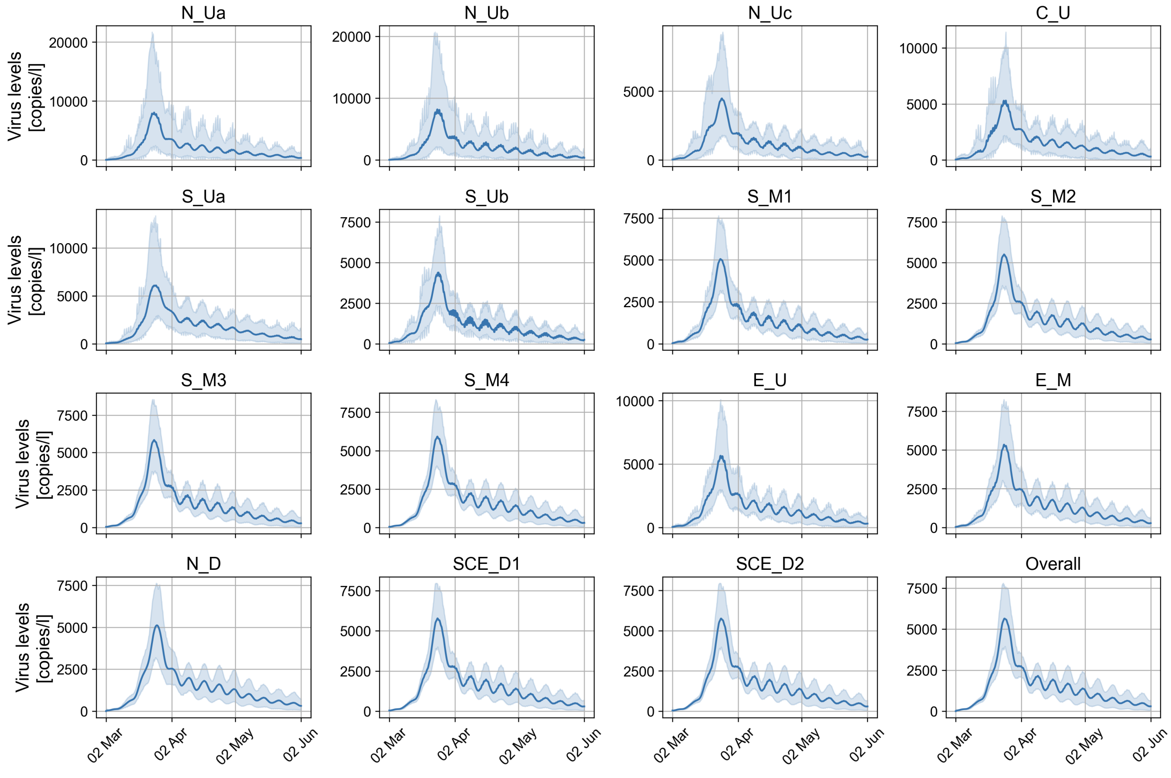


**Supplementary Figure S8: Virus levels per sampling location for the no decay & no rain scenario (uniform initialization of prevalence).** This corresponds to Scenario 1 in Supplementary Table S3.

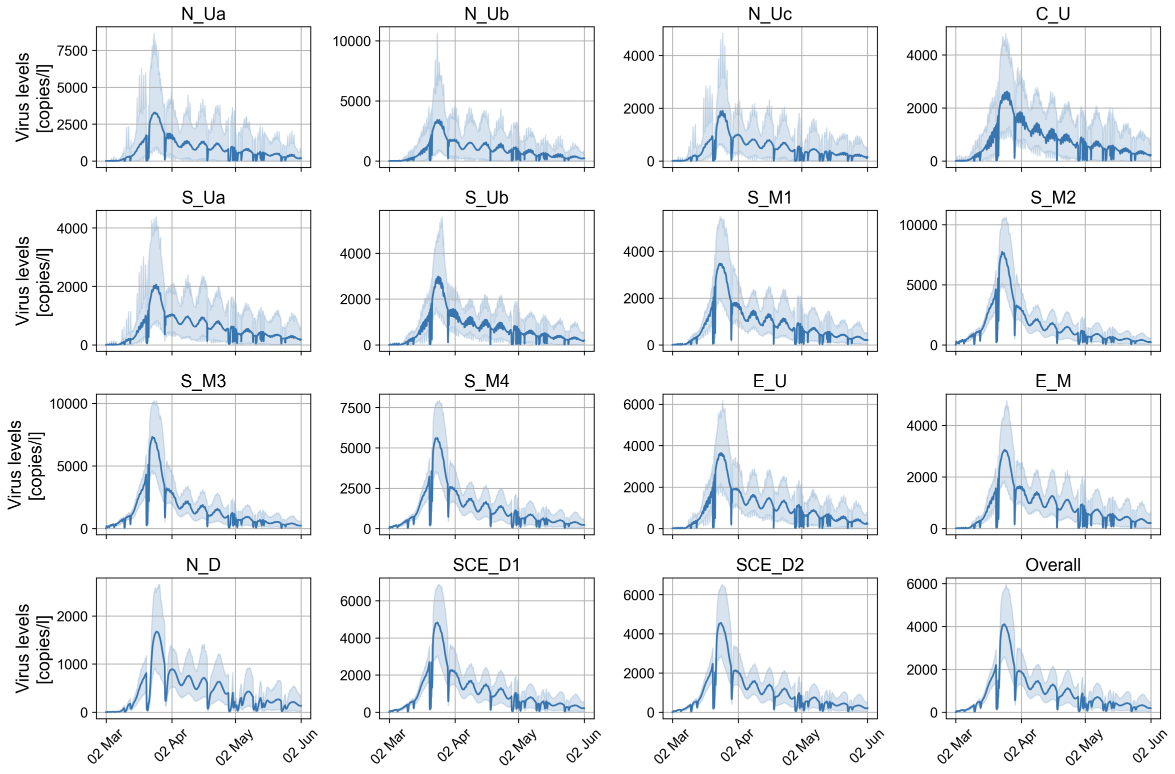


**Supplementary Figure S9: Virus levels per sampling location for the decay & rain scenario (local initialization at the city center).** This corresponds to Scenario 4 in Supplementary Table S3.

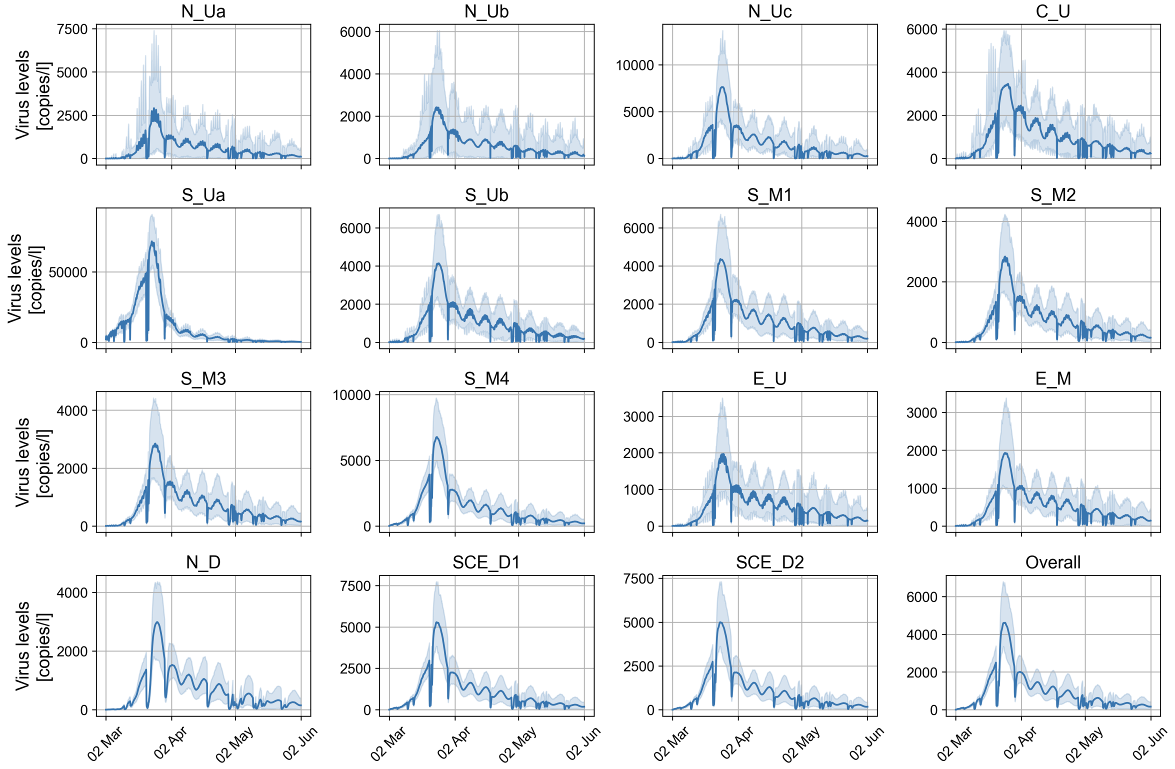


**Supplementary Figure S10: Virus levels per sampling location for the decay & rain scenario (local initialization at the city border).** This corresponds to Scenario 5 in Supplementary Table S3.

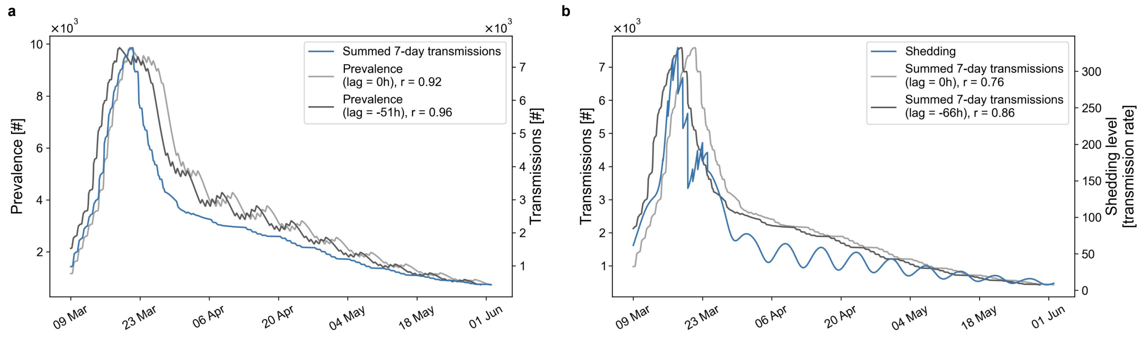


**Supplementary Figure S11: Correlation of transmissions, prevalence and respiratory shedding. a)** Mean number of transmissions summed over the last 7 days and mean prevalence over time, across 100 simulations. The prevalence shifted by -51 hours has the highest correlation with the cumulative 7-day transmissions. **b)** Mean number of transmissions summed over the last 7 days and mean respiratory shedding level over time. The cumulative 7-day transmissions shifted by -66 hours has the highest correlation with the respiratory shedding level.

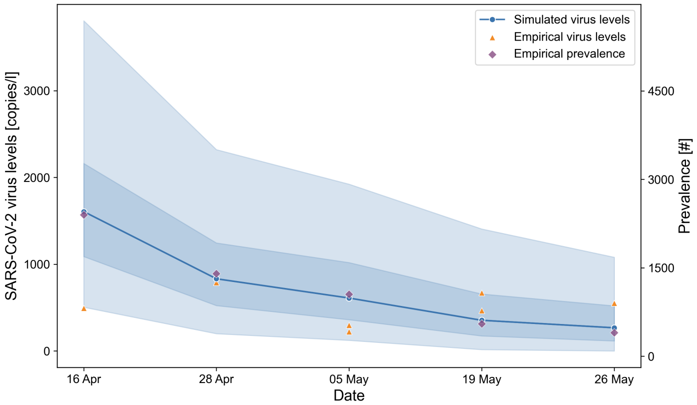


**Supplementary Figure S12: Results for fitting of RNA shedding scaling factor.** Comparison of the distribution of simulated and observed SARS-CoV-2 virus levels in wastewater across sampling locations in Munich (at each date for which at least one empirical measurement is available), based on the calibrated ABM parameters and a post-hoc estimated shedding scaling factor. The 90% interval, 50% interval, and median of the simulation results are shown. The estimated infection prevalence across Munich at each date is additionally shown in purple.
